# How do patients move within the Norwegian hospital system? A comprehensive ward- and hospital-level network analysis

**DOI:** 10.64898/2026.07.29.26359289

**Authors:** Birgitte Freiesleben de Blasio, Gianpaolo Scalia Tomba

## Abstract

Patient movements within and between hospitals create networks that can facilitate the spread of nosocomial infections. Many relevant pathogens have long-lasting carriage, allowing colonised patients to move through multiple wards across successive admissions, thereby indirectly linking wards. Yet system-wide ward-level analyses based on individual patient trajectories remain rare, even though they can reveal important features for understanding and simulating the system.

We analysed ward- and hospital-level networks using individual patient trajectories from the Norwegian Patient Registry (3.6 million registrations), covering hospital care for ∼55% of the population over one year (2012). We characterised the global network structure and, at ward level, calculated multiple centralisation measures and assessed percolation-based connectivity. From these, we identified central wards using directed K-core decomposition combined with Gaussian mixture modelling clustering. Analyses were performed separately for inpatients and all patients, and extended by linking episodes across increasing time gaps to explore how assumptions about episode continuity influence inferred connectivity. Various patient-based statistics were also analysed.

Patient movements generated sparse, regionally structured networks with clear core–periphery organisation. Inflow K-cores were larger than outflow cores, with central wards dominated by major hospital referral specialities and medical wards acting as key entry and exit points. Community structure differed by patient type: all-patient networks were largely locally contained, whereas inpatient networks spanned hospitals and regions. Temporal linking increased hub dominance only in all-patient networks, while inpatient structure remained comparatively stable. Dynamic diffusion simulations and a real outbreak analysis independently supported the identified structural backbone, demonstrating that central network hubs also represent the dominant potential pathways of patient-mediated spread under simplified transmission assumptions.

The results reveal a robust hierarchical organisational structure of patient movements that can help prioritise surveillance, while underscoring the need for pathogen-specific epidemiological and contextual data for predictive modelling.

## INTRODUCTION

Outbreaks of healthcare-associated infections (HAIs) are a persistent threat to hospital systems and patients, increasingly driven by antimicrobial-resistant (AMR) organisms that can undermine routine care (1). Transmission in healthcare settings arises from multiple interacting pathways, including healthcare worker contact, environmental persistence, and antimicrobial selection. Patient movement links wards and hospitals into a connected healthcare network, in which each transfer represents a potential pathway for pathogen dissemination. Understanding these movements is essential for anticipating how locally emerging transmission can propagate within hospitals and, in some settings, between hospitals and even regions.

Network analysis provides a natural framework for characterising patient mobility and its implications for infection spread. Previous studies have analysed patient movement at hospital (2–5), intra-hospital (6, 7), and speciality (8–10) levels to investigate healthcare connectivity and pathogen transmission. However, most have relied on aggregated flows or data from individual institutions, limiting their ability to reconstruct realised patient trajectories and characterise system-wide connectivity at ward resolution. A complementary approach (11) uses network transfer data combined with detailed patient data to estimate individual carriership probabilities, demonstrating the potential clinical applicability of such analyses.

Patient movement networks are shaped by referral hierarchies, hospital specialisation, geography, and capacity constraints. Consequently, findings from one healthcare system cannot readily be generalised to another, underscoring the need for system-specific, high-resolution data. Despite growing interest, such datasets remain rare, because individual-level patient trajectories are subject to strict data protection regulations.

The Nordic countries offer unique opportunities in this regard due to long-standing national health registries that can be linked at the individual level. Norway is a low-prevalence setting for antimicrobial-resistant pathogens, providing an opportunity to characterise structural transmission pathways as large or multi-hospital outbreaks remain uncommon.

Here, we present the first population-scale, ward-level network analysis of individual patient movements in Norway using registry data from the South-Eastern Norway Regional Health Authority. Our objectives were: (i) to characterise the organisation of patient movement networks at ward and hospital levels; (ii) identify the structural backbone of the patient movement network and classify wards according to their structural roles; and (iii) assess how alternative episode definitions influence inferred connectivity and backbone stability. We further explored patient population characteristics related to referral patterns. To aid interpretation, we complement the structural analyses with diffusion-based simulations and discuss the opportunities and limitations of registry-based patient movement data for studying nosocomial pathogen transmission.

## MATERIALS AND METHODS

### Data

We obtained individual-level data from the Norwegian Patient Registry (NPR) for the period 1 January to 31 December 2012, covering the South-Eastern Norway Regional Health Authority (HSØ), which includes the capital city of Oslo, and accounted for 55% of the national population (S1 Fig). NPR captures nearly all specialist health care activity in Norway, including public hospitals and publicly funded private providers. These data were linked with records of vancomycin-resistant *Enterococcus* (VRE) reported to the Norwegian Surveillance System for Communicable Diseases (MSIS). Ethical approval was obtained from the South-Eastern Regional Committee for Medical and Health Research Ethics (REC): 2013/1004/REK. The data were accessed for research purposes in February 2016 and July 2018.

The dataset contained information on admission and discharge dates, location (ward and hospital), age, gender, and municipality of residence for individual patients treated at all levels of care (outpatient, inpatient, and day care), and date of death for patients who died during the study period, For VRE positive patients, the linked MSIS data included the date of the first positive test and the indication for testing (clinical suspicion or screening). Each patient is uniquely identified by their Norwegian birth number, which was replaced with an anonymised ID by a third party prior to analysis to ensure privacy. The authors did not have access to information that could directly identify individuals. The dataset included 3,618,299 registrations from 416 wards across 36 hospitals and advanced treatment facilities, hereafter named hospitals (S1 Table).

### Episodes and Construction of Networks

We defined a treatment episode by aggregating all individual patient registrations with uninterrupted dates. In additional analyses, we relaxed this criterion by allowing time gaps of *d* = 1, 7, 14, 30, 60, 90, 180 and 366 days between consecutive registrations within an episode. Because the observation period spans one year, the longer time gaps are underrepresented. Relaxing episode boundaries shifts the interpretation of the network from a record of physical transfers to a representation of potential patient-mediated transmission pathways across time, while respecting the temporal order of individual patient trajectories. For example, if a patient is admitted to ward A and transferred to B during one episode, and is then readmitted *d* days later to ward A and transferred to C, the baseline definition (*d* = 0) records the transfers A → B and A → C. When the episode boundary is extended to ≥ *d* days, the patient trajectory becomes A → B → A → C, yielding the transfers A → B, B → A, and A → C, thereby indirectly linking B → C, (and also A → A with a given time lag). Of note, the epidemiological meaning of revisiting the same ward may change over time because patients, infection levels and other conditions differ; thus, temporally separated visits represent distinct contacts even when the ward label is identical. An alternative modelling choice would be to introduce direct links between non-consecutive wards visited by the same patient under extended episode definitions, in this example adding a direct link B → C. The chosen conservative approach preserves the temporal order and location-based mediation.

Based on these episodes, we constructed a 417 × 417 adjacency matrix, where each row and column represent one of the 416 wards, and an additional row and column represent transitions to and from the “outside” (e.g., home, other institutions). Each cell in the matrix indicates the frequency of patient referrals from one ward to another over the course of the year. From the adjacency matrix, we constructed a directed, weighted network. By aggregating wards within the same hospital, we further constructed a 37 × 37 adjacency matrix, representing the hospital network. This procedure was repeated separately for all registrations (*All-patient network)* and for exclusively inpatient registrations (*Inpatient network)*.

### Network analysis

The analyses were organised to characterise the patient movement networks at complementary levels: global topology, ward importance, structural robustness, backbone organisation, and community structure.

#### Network-level measures

We quantified global network organisation using density, reciprocity, and in- and out-degree centralisation (12). Degree- and strength-based assortativity were calculated to assess preferential connectivity between wards with similar numbers of connections or patient volumes (13). Observed values were compared to averages from 1,000 simulated multi-edge Erdős–Rényi networks with identical numbers of nodes and edges (no self-loops) to assess non-random structure.

#### Ward-level centrality

To characterise ward positions, we calculated in- and out-degree, in- and out-strength, closeness, betweenness, and PageRank centrality (14). Because these measures were strongly correlated and capture overlapping aspects of network position, they were treated as complementary descriptive features rather than independent indicators of ward importance.

#### Structural robustness and connectivity thresholds

Unlike conventional centrality measures, which quantify a ward’s importance within the existing network, percolation analysis quantifies how strongly a ward remains structurally embedded as network connectivity progressively deteriorates. To assess ward-level structural embeddedness, we estimated a percolation-based connectivity threshold for each ward using Monte Carlo edge-thinning. For each ward, we identified the edge-retention probability at which its strongly connected component encompassed half of all reachable wards. The same procedure was applied to weakly connected components to capture direction-independent embeddedness. Full methodological details are provided in Supporting Information,A.

#### Backbone identification and ward topology

To identify the structural backbone of patient movement, we used directed K-core decomposition (15, 16), which identifies subgroups of wards with at least *k* numbers of internal connections. Because patient flow is directional, inflow and outflow K-cores were analysed separately. Wards within the maximum K-cores were then characterised using complementary measures describing local connectivity, network position, and structural embeddedness (ward-level centrality measures and percolation connectivity threshold). Gaussian Mixture Model clustering was applied to these standardised variables to derive an interpretable typology of structural ward roles. The resulting clusters were examined in relation to ward speciality and geographic region.

#### Community structure

Community structure was identified using the Walktrap algorithm (17). The number of communities was selected by maximising modularity (Q). Community membership was used to describe the regional and speciality-based organisation of patient flow.

#### Software and visualisation

All analyses were conducted in R (RStudio 2023.12.01) using the igraph package. Network visualisations were created in Gephi 0.10.1 using the ForceAtlas2 layout.

## RESULTS

### Descriptive characteristics of patient activity

The dataset comprised over 3.6 million patient registrations during one year, of which 79% were outpatient, 15% inpatient, and 6% day-care services (Table 1). Aggregation into uninterrupted treatment episodes yielded approximately 3.3 million episodes, the majority of which involved a single ward; only 3.8% of episodes included at least one inter-ward transfer. Among inpatients, inter-ward transfers were more common, with 9.0% of episodes involving at least one transfer.

**Table 1.** Summary of patient registrations, episodes, and global ward network characteristics of the all-patient and inpatient network.

| Type | Attribute | All-patient | Inpatients |
| --- | --- | --- | --- |
| Registrations | Total, no. | 3,618,299 | 529,654 |
|  | Level of care % (inpat.,outpat.,daycare) | (14.6; 78.9; 6.5) | (100;0;0) |
| Episodes | Total, no. | 3,322,827 | 463,795 |
|  | With transfer, no. (% of total) | 127,362 (3.8%) | 41,672 (9%) |
|  | With inter-hosp. transfer, no (% of total) | 57,040 (1.8%) | 23,897 (5.1%) |
| Patients | Total, no. | 984,437 | 322,593 |
|  | Transferred, no. (% of total) | 86,020 (8.7%) | 37,463 (11.6%) |
|  | Inter-hosp. transferred, no (% of total) | 38,795 (3.9%) | 21,818 (6.8%) |
|  | Registrations per patient, median (range) | 2 (1 - 377) | 1 (1 - 138) |
|  | Episodes per patient, median (range) | 2 (1 - 258) | 1 (1 - 50) |
|  | Inter-ward movements per pat., median (range) | 0 (0 - 198) | 0 (0 - 112) |
| Ward network | Nodes, no.<sup>1</sup> | 416 | 285 |
|  | Connected | 415 | 271 |
|  | Edges, no. | 160,153 | 60,590 |
|  | Unique edges, no. | 12,626 | 5,834 |
|  | Edge density<sup>2</sup> (ER null model; p-value) | 0.07 (0.61±0.001; p<1e-3)<sup>3</sup> | 0.07 (0.53±0.001; p<1e-3)<sup>3</sup> |
|  | Reciprocity - | 0.73 (0.46±0.001; p<1e-3)<sup>3</sup> | 0.62 (0.41±0.002; p<1e-3)<sup>3</sup> |
|  | Centralised indegree - | 0.28 (0.07±0.009; p<1e-3)<sup>3</sup> | 0.27 (0.08±0.011; p<1e-3)<sup>3</sup> |
|  | Centralised outdegree - | 0.3 (0.07±0.009; p<1e-3)<sup>3</sup> | 0.27 (0.08±0.011; p<1e-3)<sup>3</sup> |
|  | Assortativity, degree<sup>4</sup> - | 0.11 (-0.002±0.002; p<1e-3)<sup>3</sup> | 0.06 (-0.003±0.003; p<1e-3)<sup>3</sup> |
|  | Assortativity, strength<sup>4</sup> - | 0.3 (0±0.003; p<1e-3)<sup>3</sup> | 0.3 (0±0.004; p<1e-3)<sup>3</sup> |

<sup>1</sup>Number of wards with registered patients at relevant level of care; i.e. in the all-patient network (inpatient, outpatient, daycare) patients, vs exclusively inpatients in the inpatient network.
<sup>2</sup>Edge density: proportion of all possible edges that are realised. For a directed network with $n$ nodes and no self-loops, network there are $n(n - 1)$ possible edges.
<sup>3</sup>Empirical p-values are obtained from Monte Carlo randomisation against the multi-edge Erdős–Rényi (ER) null model.
<sup>4</sup>Degree and strength assortativity are calculated as the Pearson correlation between source (outgoing) and target (incoming) node degree or strength across directed edges.

Approximately one-third of the region’s residents (N = 984.4K / 2785.3K) interacted with the hospital system during the year. Patient utilisation was highly uneven, with a small minority of patients accounting for a disproportionate share of registrations and episodes (S2 Table). Likewise, inter-ward transfers were concentrated among a relatively small proportion of patients, 8.7% in the all-patient network and 11.6% in the inpatient network. Most transfers were generated by patients with few recorded ward-to-ward movements.

Patient activity exhibited clear temporal patterns, with pronounced weekly and seasonal variation in registrations and transfers, including marked dips on weekends and during holiday periods (S2A, S2B, S3 Figs). Inpatient activity was less volatile over time, consistent with longer and more urgency- driven episodes of care. Also, the data showed age-specific heterogeneity in both patient counts and registrations with distinct patterns across age groups and by sex (S4 Fig).

### Global characteristics of the ward networks

The resulting patient movement networks comprised 416 wards in the all-patient network and 285 wards in the inpatient network, with annual edge weights of approximately 160K and 61K transfers, respectively (Table 1). The networks were sparse (density ≈0.07), meaning that only a small fraction of all possible ward-to-ward links were realised, consistent with a highly specialised healthcare system in which patient movements follow defined clinical pathways.

Both networks exhibited high reciprocity (0.62–0.73), indicating substantial bidirectional patient exchange between wards. In- and out-degree centralisation values (0.27–0.30) reflected a moderately hub-dominated architecture. Degree assortativity was modest but largest in the all-patient network, whereas strength assortativity was substantial and similar in both networks, indicating preferential exchange among high-volume wards. All global metrics deviated strongly from their corresponding null models (all p < 0.001), indicating a highly structured network organisation with patient exchange concentrated among interconnected wards. Visual inspection of the networks (Fig 1) highlights a pronounced Oslo-centred regional organisation.

**Fig 1.**
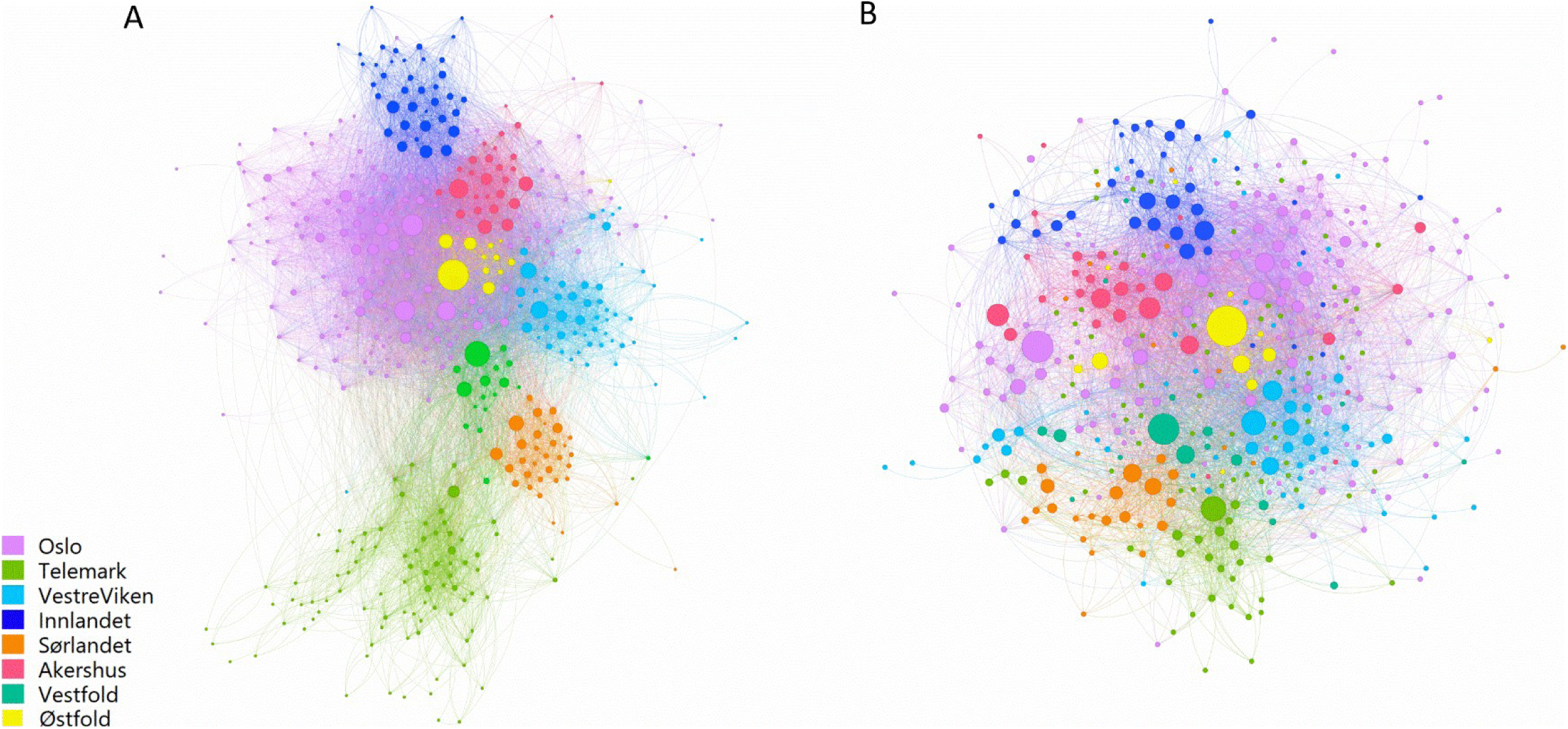
Ward-level patient transfer networks across hospitals and care contexts. (A) All patients; (B) inpatients only. Nodes represent wards and are coloured by region; node size reflects mean prevalence (average daily number of registered patients). The networks show a clear Oslo-centred structure. In the all-patient network, elongated southern “tails” suggest weaker inter-regional connectivity, whereas the inpatient network is more cohesive. Most transfers occur within regions, accounting for 80–93% of transfers in the all-patient network and 57–83% in the inpatient network (Table S3). Oslo exhibits the highest level of inter-regional exchange, highlighting its central coordinating role. Visualisation was performed in Gephi using the ForceAtlas2 layout, which places highly connected nodes centrally, less connected nodes peripherally, and clusters densely interconnected nodes.

### Ward-level characteristics

Most wards exhibited broadly balanced inter-ward patient flows, receiving and referring similar numbers of patients (Fig 2). Nevertheless, some ward types exhibited distinct referral patterns. Acute and emergency units primarily acted as sources, transferring patients onward, whereas rehabilitation wards were the clearest net recipients. Restricting the analysis to inpatients reduced overall flow volumes while preserving these broad functional patterns.

**Fig 2.**
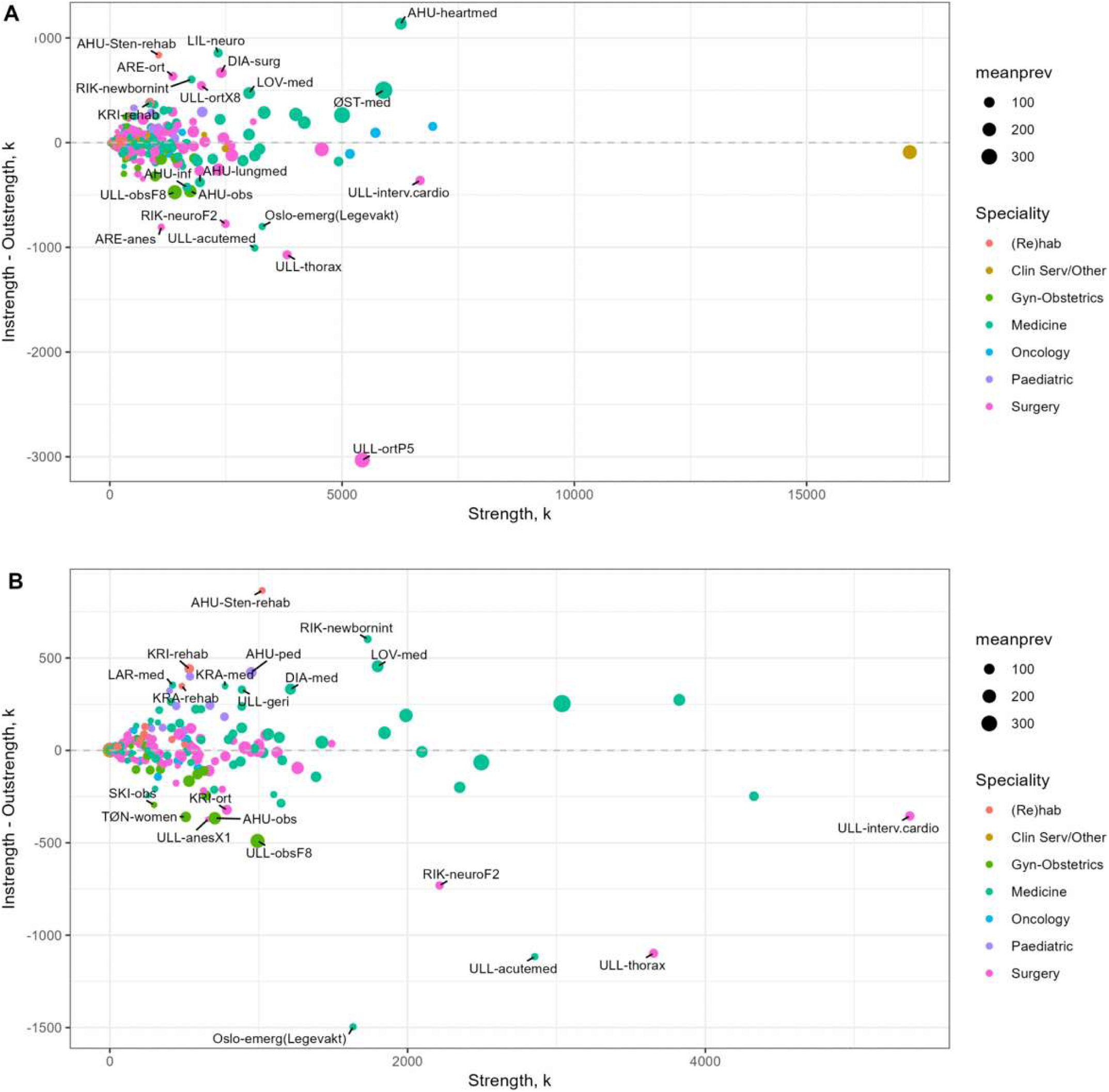
Net patient flow by ward. (A) All-patient network; (B) inpatient network. Each point represents a ward, with total patient transfer volume on the x-axis and net flow (inflow minus outflow) on the y-axis. Positive values indicate net receivers (sinks), and negative values indicate net donors (sources). The stippled line denotes neutral flow. Node size reflects mean prevalence (average daily number of registered patients), and colour indicates ward speciality. Wards with the most extreme net flow values are labelled.

Across both networks, patient flow volume and connectivity co-varied strongly (Fig 3), indicating that wards with many connections also tended to exchange large numbers of patients. However, variation between individual wards was evident: some wards combined high patient flow with only moderate connectivity, while others served as broad connectors with many links but lower throughput.

**Fig. 3.**
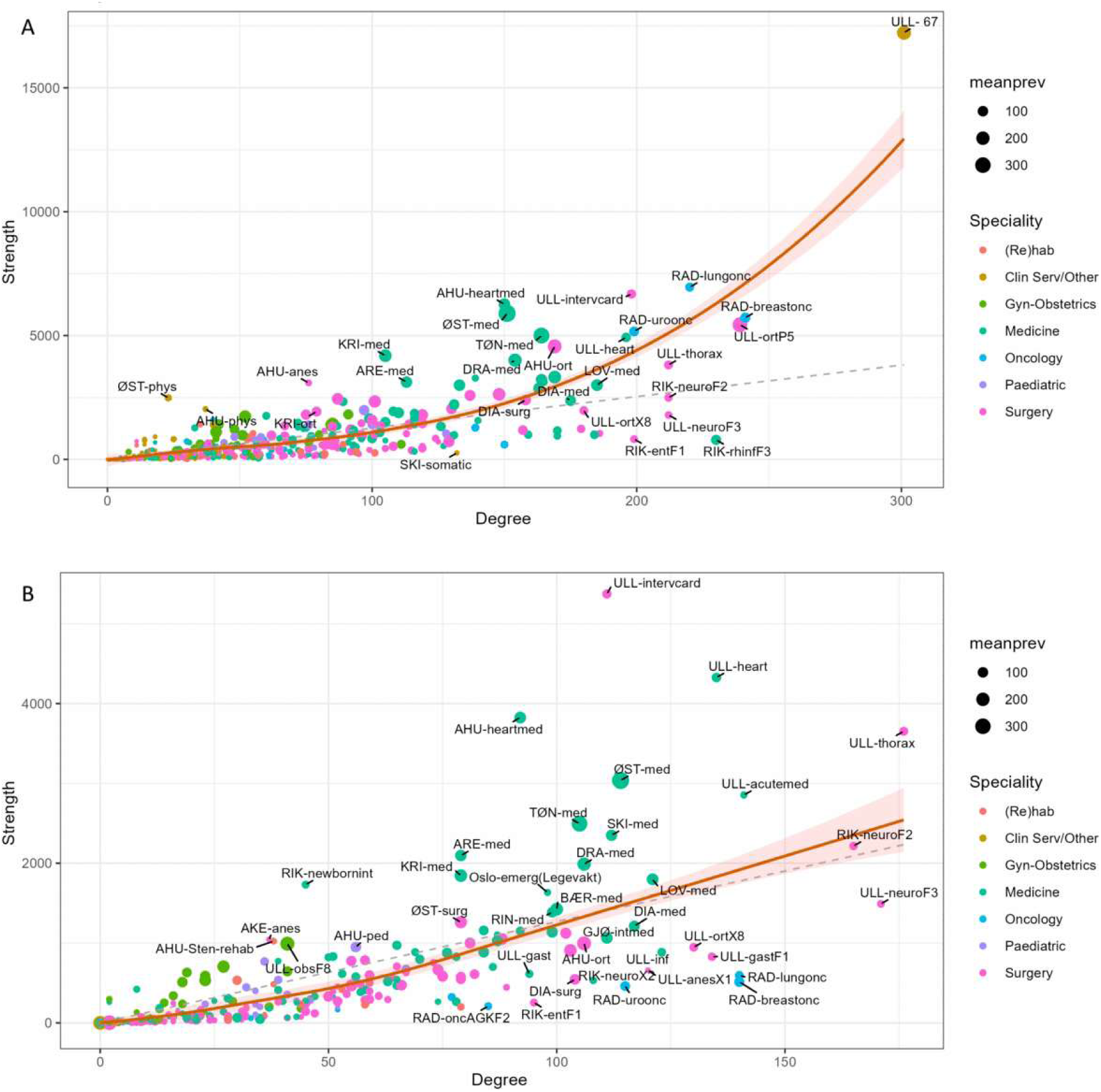
Ward-level connectivity (degree) versus total patient flow (strength). (A) All-patient network; (B) inpatient network. Node size is proportional to mean prevalence (average daily number of registered patients), and colour indicates clinical speciality. The solid red line shows a non- parametric LOESS fit, with shaded bands indicating the 95% confidence interval. The stippled line represents the expected strength under the assumption that edge weights are independent of degree (k), such that s(k) ∼ ⟨w⟩ k (24). Wards at major Oslo hospitals (Ullevål, Rikshospitalet, Radiumhospitalet) are among the most highly connected. Ullevål-67 is an extreme outlier in the all-patient network, with strong links to cancer- related services, although its specific function could not be identified. In the inpatient network, Ullevål interventional cardiology shows particularly high flow, while neurological and thoracic surgery wards are also highly connected. Medical wards at larger regional hospitals exhibit high patient flow and high prevalence but comparatively low connectivity. Selected outlier wards are labelled.

This pattern of co-variation extended to the other centrality measures (Spearman |ρ| ∼ 0.71–0.99; S5 Fig), which is expected in networks with a large, reciprocal core, where multiple alternative paths cause degree-, flow- and path-based centrality measures to converge (14, 18). The resulting redundancy motivated the use of complementary centrality measures to distinguish the structural roles of wards within the network core.

### Central K-core cluster analysis

Directed K-core decomposition identified a relatively small set of wards forming the structural backbone of patient movement, which were subsequently classified into structural roles using Gaussian mixture models (Table 2). In both networks, the inflow core encompassed a larger fraction of wards (approximately 25–30%) than the outflow core (approximately 18%), reflecting a broader structural backbone for patient intake, whereas onward transfers are concentrated through a smaller set of gateway wards.

**Table 2.**
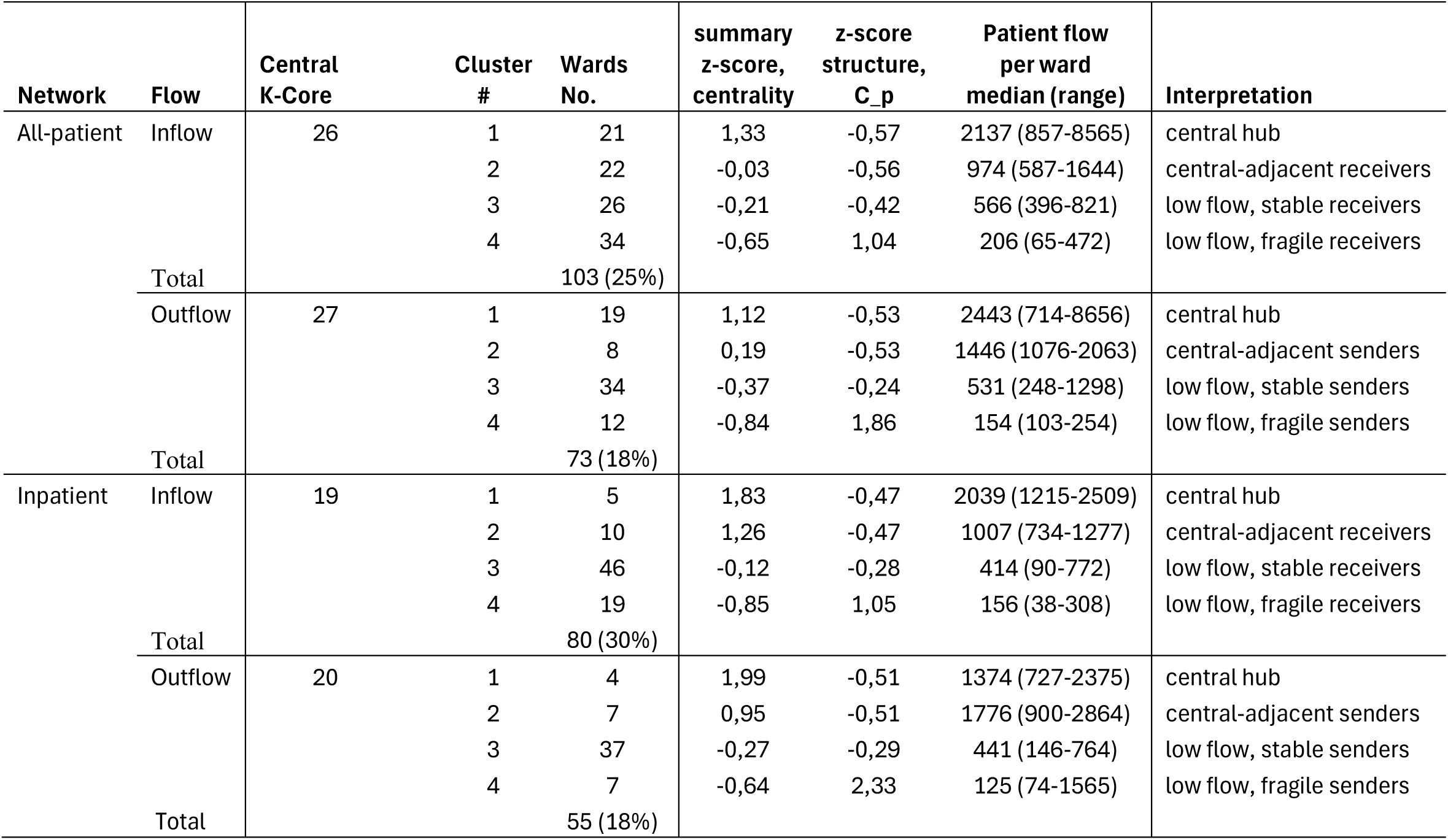
Gaussian Mixture Model (GMM) clustering of wards within central K-cores of ward-level networks. *For each network (all-patient and inpatient) and flow direction (inflow and outflow), wards in the central K-core are clustered based on scaled (z-score) multi-metric centrality and percolation thresholds. For each cluster, the table reports the K-core number, the number and proportion of wards, median annual patient flow per ward (with range), and an interpretive label describing functional role in the network. “Central hub” and “central-adjacent” clusters combine high flow and strong structural robustness; “low-flow stable” clusters are structurally robust but low in flow; and “low-flow fragile” clusters comprise sparsely connected wards that are vulnerable to fragmentation*.

The dominant hubs exhibited very high patient throughput, averaging ∼ 2000 patient transfers per ward annually, underscoring their critical role in patient movement across the hospital system.

In the inpatient network, inflow and outflow hubs overlapped substantially and were dominated by medical wards at university and major regional hospitals, and Oslo-based specialised referral units, including thoracic surgery, neurosurgery, and interventional cardiology (Fig 4). The all-patient network exhibited a similar core structure, though with a broader range of specialities represented among the hubs, including surgical and outpatient-oriented wards in cancer treatment and orthopaedics (S6 Fig).

**Fig. 4.**
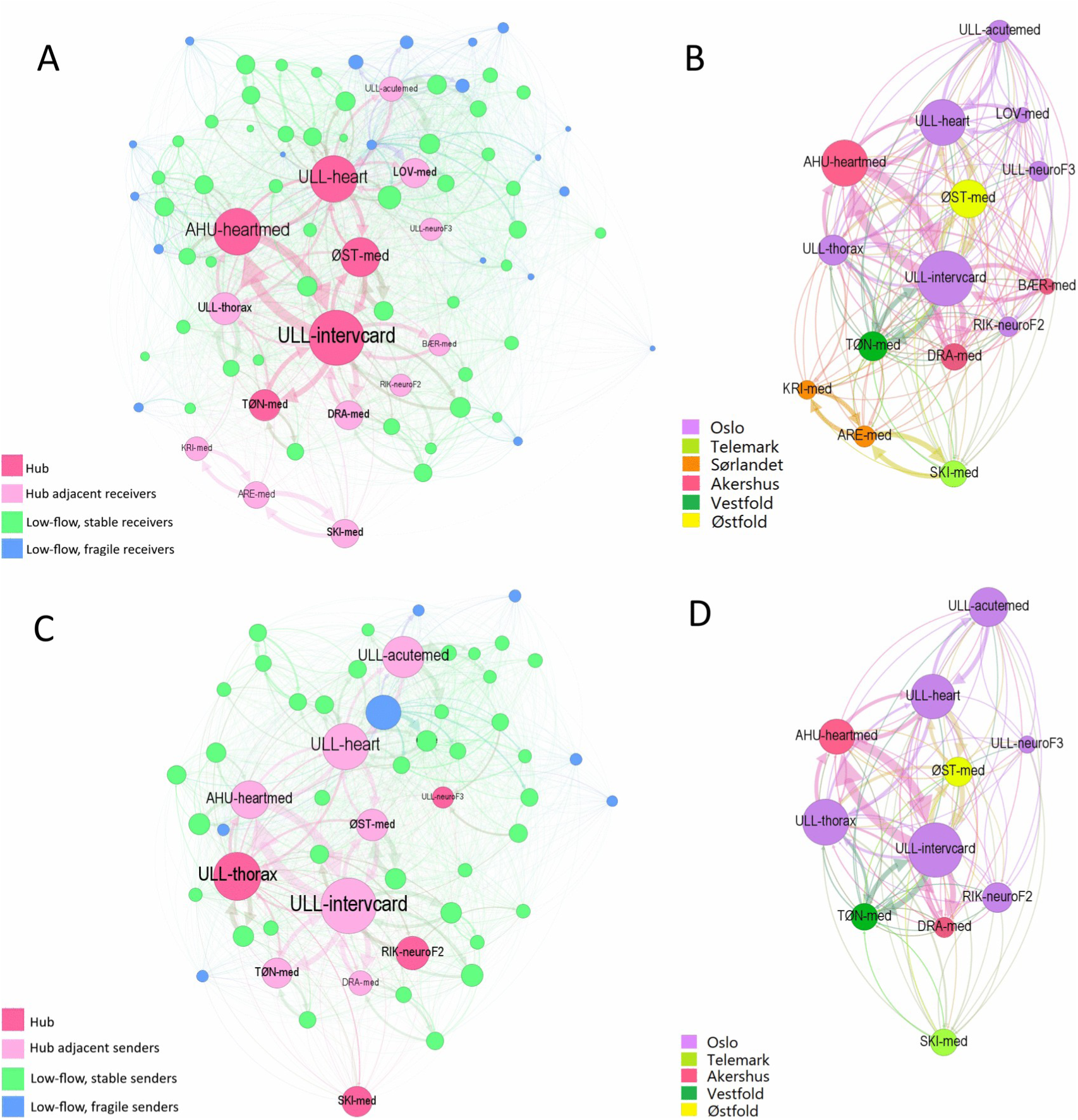
Inpatient network central K-cores: roles and regional affiliation. (A) Inflow central K-core showing GMM-derived ward roles; hub and hub–intermediary wards are labelled. (B) Hub and hub-intermediary wards in the inflow central K-core, coloured by region. (C) Outflow central K-core showing GMM-derived ward roles; hub and hub–intermediary wards are labelled. (D) Hub and hub-intermediary wards in the outflow K-core, coloured by region. Node size reflects in-strength (A–B) and out-strength (C–D). Note that wards may also maintain additional links outside the central K-core.

### Community structure

The patient movement networks exhibited strong modular organisation, with 16 multi-ward communities in the all-patient network and 9 in the inpatient network (modularity Q ≈ 0.48–0.49; Table 3). Several of the largest communities overlapped with the central K-core, confirming that highly connected wards anchor wider patterns of patient transfer.

**Table 3.**
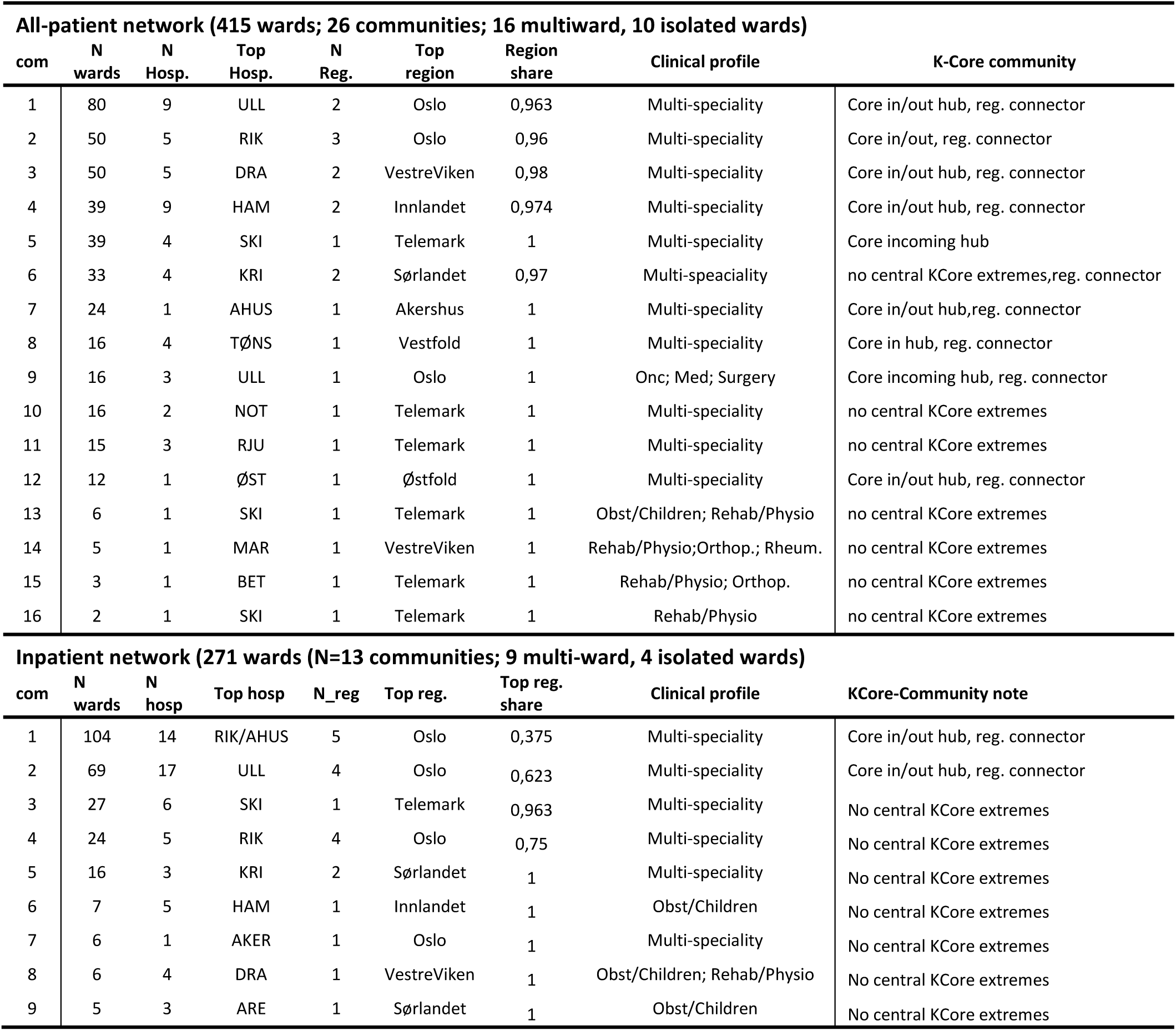
Community structure in all-patient and inpatient ward networks identified using the Walktrap community detection algorithm. *For each community, the table reports the number of wards, the number of hospitals represented, the dominant hospital and region, and the proportion of wards in the dominant region. Communities are further characterised by their clinical profiles and role within the central K-core structure*.

| **All-patient network (415 wards; 26 communities; 16 multiward, 10 isolated wards)** |  |  |  |  |  |  |  |  |
| --- | --- | --- | --- | --- | --- | --- | --- | --- |
| com | N wards | N Hosp. | Top Hosp. | N Reg. | Top region | Region share | Clinical profile | K-Core community |
| 1 | 80 | 9 | ULL | 2 | Oslo | 0,963 | Multi-speciality | Core in/out hub, reg. connector |
| 2 | 50 | 5 | RIK | 3 | Oslo | 0,96 | Multi-speciality | Core in/out, reg. connector |
| 3 | 50 | 5 | DRA | 2 | VestreViken | 0,98 | Multi-speciality | Core in/out hub, reg. connector |
| 4 | 39 | 9 | HAM | 2 | Innlandet | 0,974 | Multi-speciality | Core in/out hub, reg. connector |
| 5 | 39 | 4 | SKI | 1 | Telemark | 1 | Multi-speciality | Core incoming hub |
| 6 | 33 | 4 | KRI | 2 | Sørlandet | 0,97 | Multi-speaciality | no central KCore extremes,reg. connector |
| 7 | 24 | 1 | AHUS | 1 | Akershus | 1 | Multi-speciality | Core in/out hub,reg. connector |
| 8 | 16 | 4 | TØNS | 1 | Vestfold | 1 | Multi-speciality | Core in hub, reg. connector |
| 9 | 16 | 3 | ULL | 1 | Oslo | 1 | Onc; Med; Surgery | Core incoming hub, reg. connector |
| 10 | 16 | 2 | NOT | 1 | Telemark | 1 | Multi-speciality | no central KCore extremes |
| 11 | 15 | 3 | RJU | 1 | Telemark | 1 | Multi-speciality | no central KCore extremes |
| 12 | 12 | 1 | ØST | 1 | Østfold | 1 | Multi-speciality | Core in/out hub, reg. connector |
| 13 | 6 | 1 | SKI | 1 | Telemark | 1 | Obst/Children; Rehab/Physio | no central KCore extremes |
| 14 | 5 | 1 | MAR | 1 | VestreViken | 1 | Rehab/Physio;Orthop.; Rheum. | no central KCore extremes |
| 15 | 3 | 1 | BET | 1 | Telemark | 1 | Rehab/Physio; Orthop. | no central KCore extremes |
| 16 | 2 | 1 | SKI | 1 | Telemark | 1 | Rehab/Physio | no central KCore extremes |

| **Inpatient network (271 wards (N=13 communities; 9 multi-ward, 4 isolated wards)** |  |  |  |  |  |  |  |  |
| --- | --- | --- | --- | --- | --- | --- | --- | --- |
| com | N wards | N hosp | Top hosp | N_reg | Top reg. | Top reg. share | Clinical profile | KCore-Community note |
| 1 | 104 | 14 | RIK/AHUS | 5 | Oslo | 0,375 | Multi-speciality | Core in/out hub, reg. connector |
| 2 | 69 | 17 | ULL | 4 | Oslo | 0,623 | Multi-speciality | Core in/out hub, reg. connector |
| 3 | 27 | 6 | SKI | 1 | Telemark | 0,963 | Multi-speciality | No central KCore extremes |
| 4 | 24 | 5 | RIK | 4 | Oslo | 0,75 | Multi-speciality | No central KCore extremes |
| 5 | 16 | 3 | KRI | 2 | Sørlandet | 1 | Multi-speciality | No central KCore extremes |
| 6 | 7 | 5 | HAM | 1 | Innlandet | 1 | Obst/Children | No central KCore extremes |
| 7 | 6 | 1 | AKER | 1 | Oslo | 1 | Multi-speciality | No central KCore extremes |
| 8 | 6 | 4 | DRA | 1 | VestreViken | 1 | Obst/Children; Rehab/Physio | No central KCore extremes |
| 9 | 5 | 3 | ARE | 1 | Sørlandet | 1 | Obst/Children | No central KCore extremes |

In the all-patient network, only 31% of communities were multi-regional (S6 Table), and these were generally dominated by a single region. In contrast, the inpatient network contained a larger proportion of multi-regional communities (44%; S6 Table) with a more balanced regional composition, consistent with referral-based pathways centred on major hospitals in the Oslo region. These findings suggest that inpatient transfers are the principal structural links between otherwise regionally organised hospital networks.

A few distinct speciality-specific communities also emerged. In the all-patient network, rehabilitation and oncology formed separate communities, the latter centred on specialised cancer services in Oslo. In the inpatient network, distinct modules of obstetric and paediatric services likewise formed separate communities.

### Episode time delay sensitivity

Patient-movement networks derived from administrative episodes implicitly assume that connectivity arises only within uninterrupted episodes. However, many healthcare-associated pathogens have prolonged carriage states, such that undetected colonised patients may traverse multiple wards across successive admissions, creating epidemiological links (“memory”) between wards that are not directly connected through administrative episodes.

Allowing a time gap between consecutive discharge and admission registrations to be treated as the same episode substantially increased the inferred number of ward-to-ward movements, up to ∼six-fold (all patients) and two-fold (inpatients). Most of these additional movements occurred within the first one to three months between administrative episodes with little increase thereafter (Fig 5), consistent with the strongly right-skewed empirical distribution of readmission intervals (S7 Fig). However, these patterns should be interpreted with caution because the one-year observation period increasingly truncates long inter-admission intervals toward the end of the calendar year, as also reflected in the cumulative incidence of readmission (S8–S9 Figs).

**Fig. 5.**
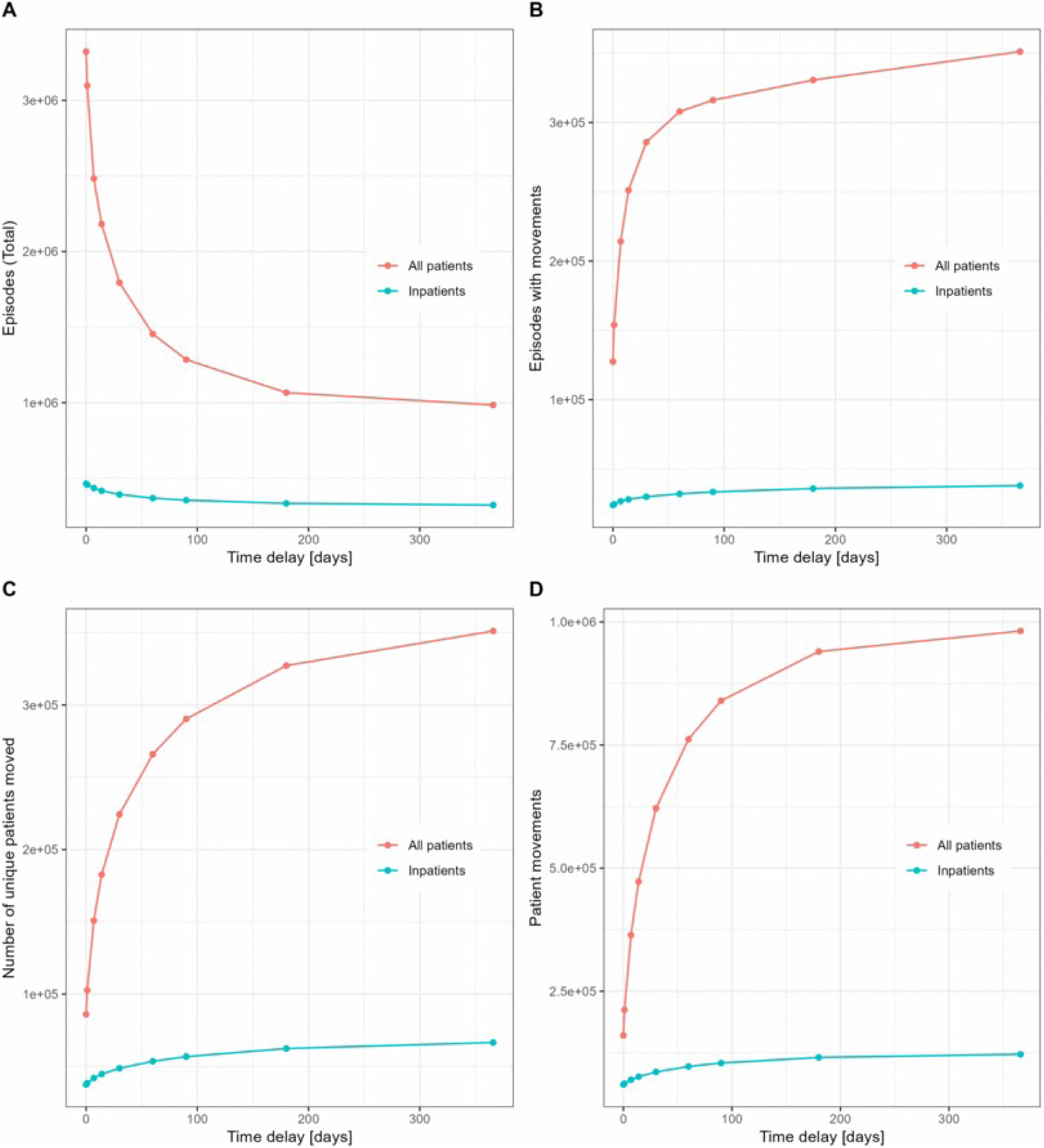
Effect of the episode-linking temporal window on inferred ward-to-ward patient movements. (A) Total number of admission episodes; (B) Number of episodes with ward-to-ward movements; (C) Number of patients experiencing ward-to-ward movements; (D) Number of inferred ward-to-ward movements. The x-axis shows the temporal linking window (days) used to connect consecutive admission episodes for the same patient. Increasing the window links temporally proximate episodes into continuous trajectories, allowing indirect ward-to-ward connections across intermediate community (“outside”) stays. Red lines indicate all patients and teal lines indicate inpatients. Most changes occur at short linking windows, with diminishing effects as the window increases; however, these patterns may be influenced by truncation of longer inter-episode intervals due to the one-year observation period.

Temporal linking significantly densified the networks, but its structural consequences differed by patient type. In the all-patient network, increased connectivity was accompanied by greater hub dominance and declining assortativity (S4 Table), reflecting temporally clustered outpatient activity feeding into major hubs. In contrast, the inpatient network largely preserved its centralisation and assortative structure, indicating a more cohesive and temporally stable topology. The identity of central backbone wards remained stable, especially among inpatients, even as peripheral connectivity increased (S5 Table), while cross-regional mixing declined for both networks (S6 Table).

To distinguish the effect of temporal aggregation from purely structural connectivity, we compared random edge thinning with analyses that preserve the observed temporal ordering of patient movements. Random edge thinning produced lower percolation connectivity thresholds than analyses that preserved the calendar time of patient movement sequences. In the baseline case (d = 0), the weakly connected component percolated at retentions of approximately 1 and 1.8 structural “days” in the all-patient and inpatient networks, respectively, whereas preserving the temporal sequences of movements required roughly 2 days (S7-S8 Tables).

The sensitivity of the percolation threshold to the maximum allowed registration time gap varied across networks. In the all-patient network, increasing the gap from 0 to 90 days reduced the percolation threshold by approximately an order of magnitude under random thinning, compared with approximately a four-fold reduction when the temporal order or patient movements was preserved. In contrast, the inpatient network showed a much smaller response, with the threshold decreasing by only about half and remaining close to the calendar-respecting estimate. Inpatient referrals were more evenly distributed over time, making random edge thinning a reasonable approximation of instantaneous network structure. Outpatient and day-care activity was more temporally clustered, causing annual aggregation to overestimate short-term connectivity in the all-patient network. Strongly connected components (bidirectional) had higher percolation thresholds overall but exhibited the same qualitative response to increasing episode-linking windows.

### Aggregated hospital networks

To provide an administratively interpretable perspective and assess the robustness of ward-level findings to aggregation, we collapsed the ward networks to the hospital level. Hospitals differ substantially in how finely care is subdivided into wards, a heterogeneity that can inflate the apparent importance of individual wards in ward-level analyses. Aggregation reduces this distortion while preserving the main patient-movement channels between institutions and regions. Fig 6 shows the resulting hospital networks for all patients and inpatients, including intra-hospital movements and the central inflow K-core. In both networks, strong within-region connectivity is evident, organised around an Oslo-centred backbone. Ullevål Hospital, the largest hospital in Oslo, emerges as a key regional hub bridging multiple regions.

**Fig. 6.**
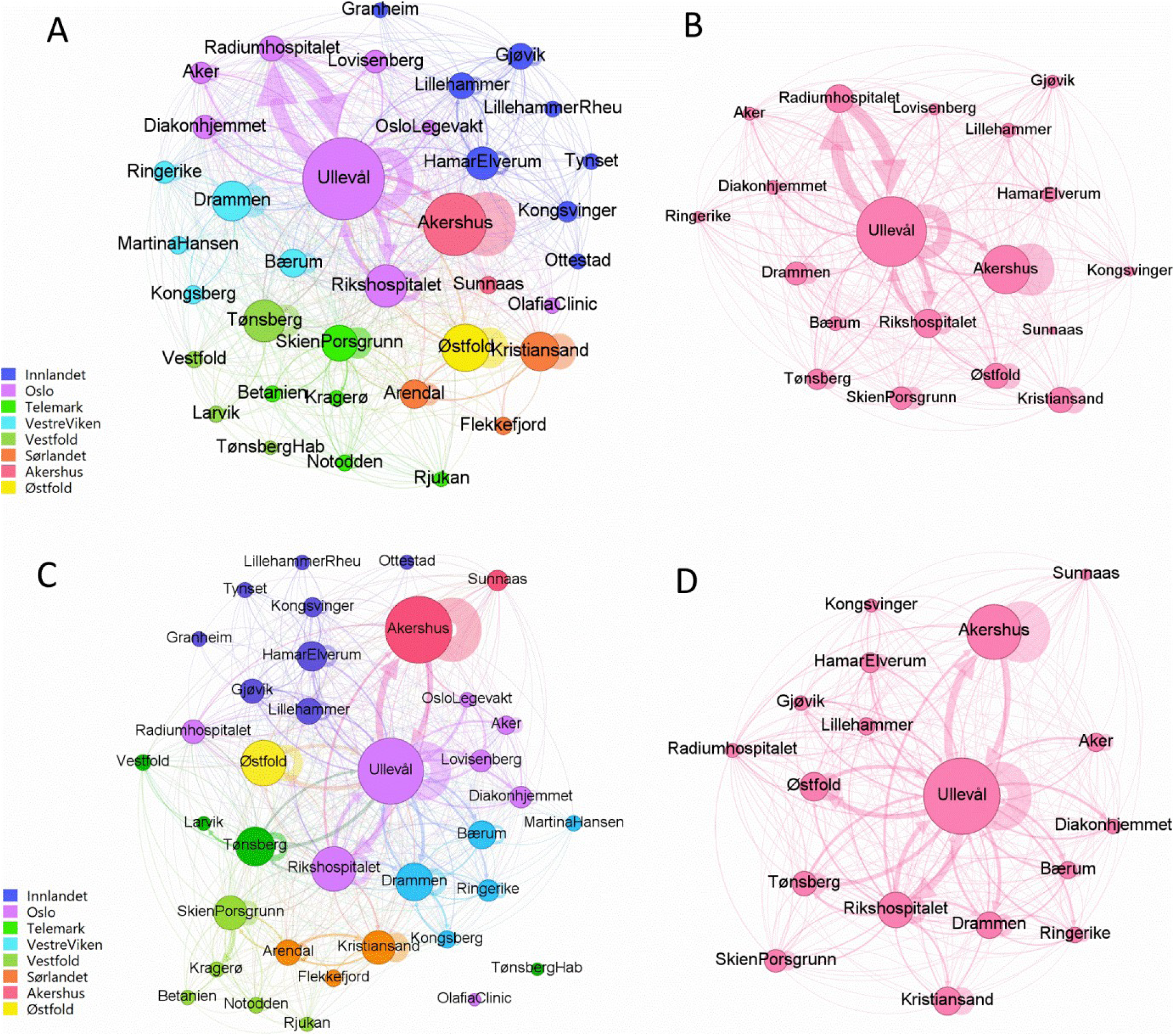
Aggregated hospital networks. (A) All-patient network, with node size scaled to mean daily prevalence (average number of admitted patients per day) and nodes coloured by region; (B) Inflow central K-core of the all-patient network (K = 19 hospitals); (C) Inpatient network, with node size scaled to mean daily prevalence and nodes coloured by region; (D) Inflow central K-core of the inpatient network (K = 18 hospitals). Self-loops represent internal patient transfers within the same hospital. Two hospitals—Tønsberg Habilitation and the Olafia Clinic (a specialised STI treatment clinic)—were isolated in the inpatient network. The outflow and inflow central K-cores were highly similar, sharing 17 hospitals (17/21 in the all-patient network and 17/18 in the inpatient network; see S6 Fig).

The hospital-level networks also enable a direct comparison of intra- and inter-hospital movements. Among all patients, movements to and from outside the hospital system dominate (95.40%), while intra-hospital transfers were slightly more frequent than inter-hospital transfers (2.51% vs 2.08%). Among inpatients, inter-hospital transfers accounted for a larger share than intra-hospital movements (6.59% vs 4.96%). Hospital-level medians and interquartile ranges were broadly consistent with these system-level values, though variability was higher in the inpatient network (S9 Table). Overall, aggregation to the hospital level preserved the Oslo-centred backbone and regional referral structure identified in the ward-level analyses despite differences in organisational granularity.

## Discussion

We analysed a detailed, population-based dataset of ward-level patient movements across the Norwegian hospital system, capturing care for over half of the national population in a single year. Our findings show that patient movements form sparse, highly non-random, regionally structured networks. These networks are organised around a structural backbone that sustains connectivity at both ward and hospital levels.

This work was undertaken to better characterise how nosocomial infections can spread within and between hospitals and to provide a realistic basis for mathematical modelling of intra- and inter-hospital transmission dynamics (19). Understanding transmission dynamics within hospitals is complex; patient movements constitute only one part of a multifaceted system that also includes healthcare worker contacts, environmental persistence, infection prevention practices, and the underlying susceptibility and infectivity of different patient populations.

A key strength of this study is the use of individually linked patient trajectories at ward resolution, providing complementary insights into inpatient dynamics embedded within the wider all-patient flow. Such datasets remain rare: a 2026 systematic review identified only two of 79 studies with individually linked, ward-level data across healthcare systems (20). Individual-level patient trajectories allowed us to assess whether connectivity inferred from aggregated movement data remains robust after accounting for the temporal ordering and dispersion of patient movements.

Previous patient-movement studies have primarily relied on individual centrality measures or global network metrics to identify important wards or hospitals. While K-core decomposition and centrality measures are widely used in network analysis (21, 22), their combined use with probabilistic clustering has not, to our knowledge, been applied to identify structural roles in system-wide patient-movement networks. Our analysis provides several insights relevant to nosocomial infection spread by showing which wards predominantly receive, redistribute and bridge patient flow:

*(i) Most patients arrive from outside the hospital system*. In our data, patient inflow from the community is large, while ward-to-ward transfers are low (≈5–12%), making the risk of ward introduction sensitive to community prevalence. This underscores the importance of monitoring community indicators (e.g., positivity rates, outbreak signals) and scaling up admission screening during periods of elevated risk.
*(ii) Most patient movements are local, but inpatients bridge hospitals*. Most ward-to-ward transfers occur within the same hospital, indicating that opportunities for onward spread are predominantly local and therefore most relevant to routine outbreak investigations. Inpatient transfers, however, more frequently cross hospital boundaries, creating a more cohesive referral network that links specialised services across regions. In contrast, including outpatient and day-care episodes, which account for most patient registrations, yields a network that is more regionally confined and dominated by local service structures.
*(iii) Different clinical specialities have distinct roles within the structural backbone.* The stable structural backbone, particularly in the inpatient network, revealed distinct roles for different clinical specialities. Medical wards consistently function as hospital hubs, receiving patients from multiple clinical pathways and acting as key transfer nodes for patient movement both within and between hospitals. From a structural perspective, these wards may therefore represent priorities for surveillance. In contrast, specialised surgical wards, including thoracic surgery, neurosurgery, and interventional cardiology, function mainly as redistribution hubs, transferring patients onwards to a wide range of wards across regional referral pathways.
*(iv) Temporal linking of episodes introduces latent connectivity that may substantially alter network topology.* Apparent connectivity depended strongly on the permitted gap between registrations within episodes; most extra links arise when merging episodes within 1–3 months, consistent with the carriage durations of many pathogens relevant to nosocomial transmission. Temporal linking densifies networks; however, the inpatient network largely preserves its core architecture, whereas the all-patient network, which predominantly bridges outpatient activity, yields a more hub-dominated yet regionally confined structure.

Time is a critical dimension for interpreting patient-movement networks. Hospital transmission unfolds on an instantaneous network, whereas our analyses rely on annual aggregates. Random edge thinning provides a heuristic approximation to shorter observation windows, although this interpretation depends on strong stationarity assumptions. Our analyses revealed pronounced temporal heterogeneity, implying that annual aggregation may overestimate short-term connectivity. This distinction is critical for outbreak response, where rapid structural reachability may overstate near-term transmission risk if temporal constraints are ignored.

Alternatively, random edge thinning can be interpreted as a system-wide, scaled reduction in patient movements, or in the risk of patients carrying pathogens. In our reciprocal, core–periphery networks, such uniform dilution barely disrupts connectivity until very high removal levels because redundant routes through the dense core are preserved.

So how and when can registry-based patient movement data be used in modelling, and what can they reveal? One application is to incorporate patient movement data into network diffusion models. Simulations of a ward-level SI model parameterised by observed patient volumes (Supporting Information, B) generated spread that preferentially followed the dense core and high-connectivity corridors. Wards reached earliest closely corresponded to those identified by the K-core and GMM analyses, suggesting that the structural backbone captures the principal diffusion pathways. This correspondence is not evident from the K-core analysis alone, as wards with high patient flow or centrality may lie outside the maximum K-core. In this stylised setting, targeted edge- and node-based interventions further illustrate how structural information can be used to qualitatively compare intervention scenarios and identify candidate intervention targets. A vancomycin-resistant *Enterococcus* outbreak at Østfold Hospital (23) provided a practical illustration (Supporting Information, C). Patient-flow rankings identified the most affected wards in the outbreak, while sporadic cases at other hospitals did not align with the dominant system-wide movement pathways. Together, these examples illustrate how registry-based patient movement data can generate hypotheses about potential transmission pathways, support the prioritisation of surveillance and outbreak investigations, and inform the design of more detailed pathogen-specific models.

Moving from structural insight to epidemiologic realism, however, requires additional local data and knowledge: pathogen-specific parameters (colonisation duration, shedding, dose–response), ward-level hazards (e.g., plumbing/equipment/procedure issues), staff movements, time-stamped infection prevention and control (IPC) actions, and patient-specific susceptibility and transmissibility in various settings. Although we had access to the main ICD-10 diagnosis for each treatment episode, we lack an empirically justified mapping from diagnosis categories and ward types to acquisition, carriage and transmission risk. While it is straightforward to simulate hypothetical “what-if” scenarios given simplifying assumptions, such model outputs are inherently speculative without data to anchor them. Much of the required information already exists at the hospital level but must be systematically collected and combined with movement data to enable national- or regional-scale inference.

Furthermore, interpreting hospital outbreak and prevalence data is challenging because screening is typically triggered by case detection, such as a severe infection. Therefore, the observed prevalence reflects both the emerging outbreak and previously undetected background carriage, which are difficult to disentangle. Even admission screening data are commonly biased, as they often target specific patient groups, e.g. patients with a travel history to high-prevalence settings. Estimating transmission probabilities therefore requires careful consideration of the sampling process that generated the data.

The Norwegian setting offers particular opportunities to validate this framework. Norway is a low-prevalence country for resistant pathogens, and large-scale multi-hospital outbreaks are uncommon. This enables detailed investigation of transmission routes within hospitals by combining patient transfer data with microbiology, electronic health records, and whole-genome sequencing. The low background prevalence also makes it easier to detect local environmental or ward-specific effects that may otherwise be obscured in high-prevalence settings. Future work should integrate hospital patient movements between hospitals and nursing homes, which may serve as important gateways for pathogen introduction into and reintroduction from the hospital system.

This study has several limitations. First, ward granularity varied between hospitals, reflecting local administrative and organisational practices. Administrative wards typically cover several sub-units and may span physically separated buildings. While ward-level aggregation is appropriate for analysing system-wide patient flow, this heterogeneity may influence the apparent importance of individual wards. Second, the analysis is subnational, introducing boundary censoring of patient trajectories and potentially underestimating cross-region connectivity, although only a small fraction of patients (∼3-4%) treated in HSØ hospitals resided outside the covered counties. The study covers a single year, precluding assessment of multi-year dynamics and seasonal persistence, while the admission-delay distribution is biased toward shorter delays. Finally, the structural role classifications are based on a Gaussian mixture model and should be viewed as one plausible partition of the underlying network characteristics. Alternative clustering approaches may yield somewhat different group assignments of wards, particularly for wards with intermediate characteristics, although the broader patterns of structural organisation are unlikely to be materially affected.

In conclusion, registry-based patient movement data provide a powerful lens through which to characterise the organisation of hospital systems and identify the principal pathways connecting wards and hospitals. Translating these structural insights into epidemiological understanding and predictive modelling, however, requires integration with pathogen-specific data and local epidemiological information.

## Data Availability

The data used in this study are not publicly available because they contain individual-level health information protected under Norwegian legislation and the General Data Protection Regulation (GDPR). Researchers who fulfil the legal and ethical requirements for access may apply for access through the Norwegian Health Data Service via http://www.helsedata.no.

## ACKNOWLEDGEMENTS

The authors would like to thank Petter Elstrøm, Ragnhild Raastad and Jørgen Bjørnholt at the Norwegian Institute of Public Health for valuable discussions during the development of this work.

## FUNDING

BFdB was funded by NordForsk, (https://www.nordforsk.org/), grant number 209400 and grant number 200568. The funder played no role in the study design, data collection and analysis, decision to publish or preparation of the manuscript.

## Notes

### Competing Interest Statement

The authors have declared no competing interest.

### Author Declarations

South-Eastern Regional Committee for Medical and Health Research Ethics, Norway, gave ethical approval for this work (2013/1004/REK).

### Summary of Updates

Added Supporting Information and metafiles

