## Supporting Information for "How do patients move within the Norwegian hospital system? A comprehensive ward- and hospital-level network analysis"

#### A. Ward-level structural connectivity threshold (percolation-based)

Percolation on networks provides a general framework for quantifying structural robustness and connectivity thresholds (1-3). In bond percolation, edges are progressively removed at random, and the critical percolation threshold, $C_p$, denotes the minimum fraction of retained edges required to sustain large-scale network connectivity.

Classical percolation theory characterises a global network property through the emergence of a giant connected component. Here, our objective instead was to characterise the structural embeddedness of individual wards. We therefore adapted the percolation framework to estimate ward-specific connectivity thresholds, defined as the edge-retention probability at which an individual ward becomes connected to a predefined proportion of the hospital network. Bond percolation was applied to the directed ward-level patient-movement networks aggregated over one year, analysing inpatient and all-patient networks separately. Connectivity was evaluated under both weak and strong definitions. Weak connectivity ignores edge direction and therefore captures hierarchical or acyclic patient flows (e.g. ICU → general ward → discharge), whereas strong connectivity requires mutual reachability via directed paths and therefore represents fully bidirectional patient transfer.

##### Estimation of bond percolation threshold

For the ward-level analysis, percolation thresholds were estimated on the network obtained from the annual adjacency matrix, restricted to the giant weakly connected component (isolated wards were excluded). This baseline set of $N_W$ wards was used in all subsequent connectivity calculations.

Let $G = (V, E)$ denote the resultant directed network with $|V| = N_W$. To implement bond percolation, each edge $e \in E$ was assigned an independent random number

$$u_e \sim \text{Uniform}(0,1)$$

For a given edge-retention probability $p$, an edge was retained whenever $u_e \leq p$, yielding a graph in which a fraction $p$ of edges is kept. The same edge-specific random values were reused across all values of $p$, producing nested edge-thinned networks in which increasing $p$ only adds edges.

For a given edge-thinned network and a ward $i$, connectivity was quantified as the size of the weakly or strongly connected component containing $i$, normalised by $N_W$. This defines a ward-level connectivity fraction

$$f_i(p) = \frac{|C_i(p)|}{N_w}; \qquad f_i(p) \in [0,1]$$

where $C_i(p)$ denotes the connected component containing ward $i$ at edge-retention probability $p$.

Connectivity was evaluated over a logarithmically spaced grid of edge-retention probabilities spanning $10^{-5}$ to 1. For each simulation replicate, edge-specific random values were generated once and reused across all $p$-values to preserve nestedness. Ward-specific connectivity fractions $f_i(p)$ were then evaluated for every value of $p$.

For each ward, $i$, we defined the ward-specific percolation threshold $\hat{C}_p(i)$ as the value of $p$ at which $f_i(p) \approx 0.5$, i.e. the connected component including ward $i$ reaches 50% of the wards in the giant connected component. Low thresholds indicate well-embedded wards that become integrated into the broader network at low edge retention levels, whereas high thresholds indicate structurally peripheral wards. The specific choice of a 50% percolation threshold is not of primary interest. Rather, it provides a consistent ranking of wards according to their structural embeddedness. Similar rankings were obtained using thresholds of 25% and 75%, indicating that conclusions are robust to the chosen cutoff.

For the nested networks, replicate-specific estimates $\hat{C}_p^j(i)$were obtained by linearly interpolating $f_i(p)$ between the first $p_k$ where $f_i(p_k) > 0.5$ and the previous grid point $p_{k-1}$. Ward-specific thresholds were summarised across 1,500 simulation replicates using the mean, median and interquartile range. As a network-level measure of structural robustness, we summarised the distribution of the earliest ward-specific threshold (the minimum across wards within each simulation replicate).

Full ward-level percolation profiles were also computed to describe how connectivity evolves as a function of $p$. The profiles characterise the evolution of ward connectivity across the full range of edge-retention probabilities (Fig. SuppA.1), displaying a characteristic bi-modal distribution of $C_i(p)$-values for intermediary p-values (Fig.SuppA.2)

The percolation thresholds of ward-level strongly connected components were used in the GMM clustering procedure, alongside ward-specific centrality measures, to identify wards occupying distinct structural roles within the maximum K-cores. The corresponding network-level measures (minimum ward-specific thresholds) for the weakly and strongly connected components were used to quantify changes in structural robustness when relaxing the allowed time gap between discharge and readmission for individual patients.

Supplementary S1 Movie and S2 Movies illustrate this process dynamically for representative central, intermediate and peripheral wards.

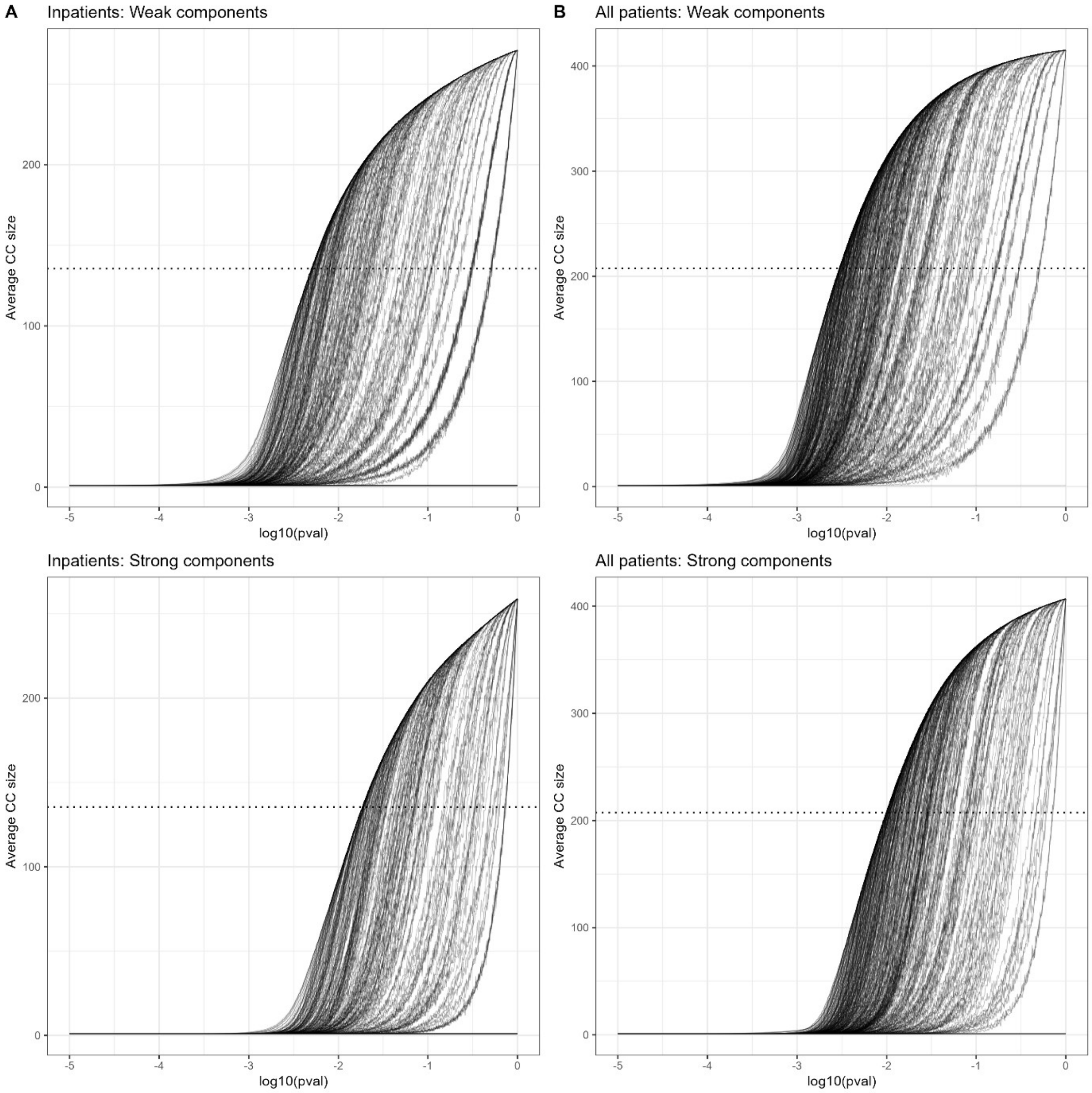


**Fig SuppA.1: Ward-specific average connected component size as a function of edge-retention probability, p-val.** Each curve shows the average number of wards reachable from a given ward as a function of the edge-retention probability (on the x-axis, shown on a $\log_{10}$ scale), averaged over simulations. Top panels show weakly connected components and bottom panels show strongly connected components for the inpatient network (A, left panel) and the all-patients network (B, right panel). The horizontal dotted lines indicate the 50% connectivity threshold used when estimating $\hat{C}_p(i)$.

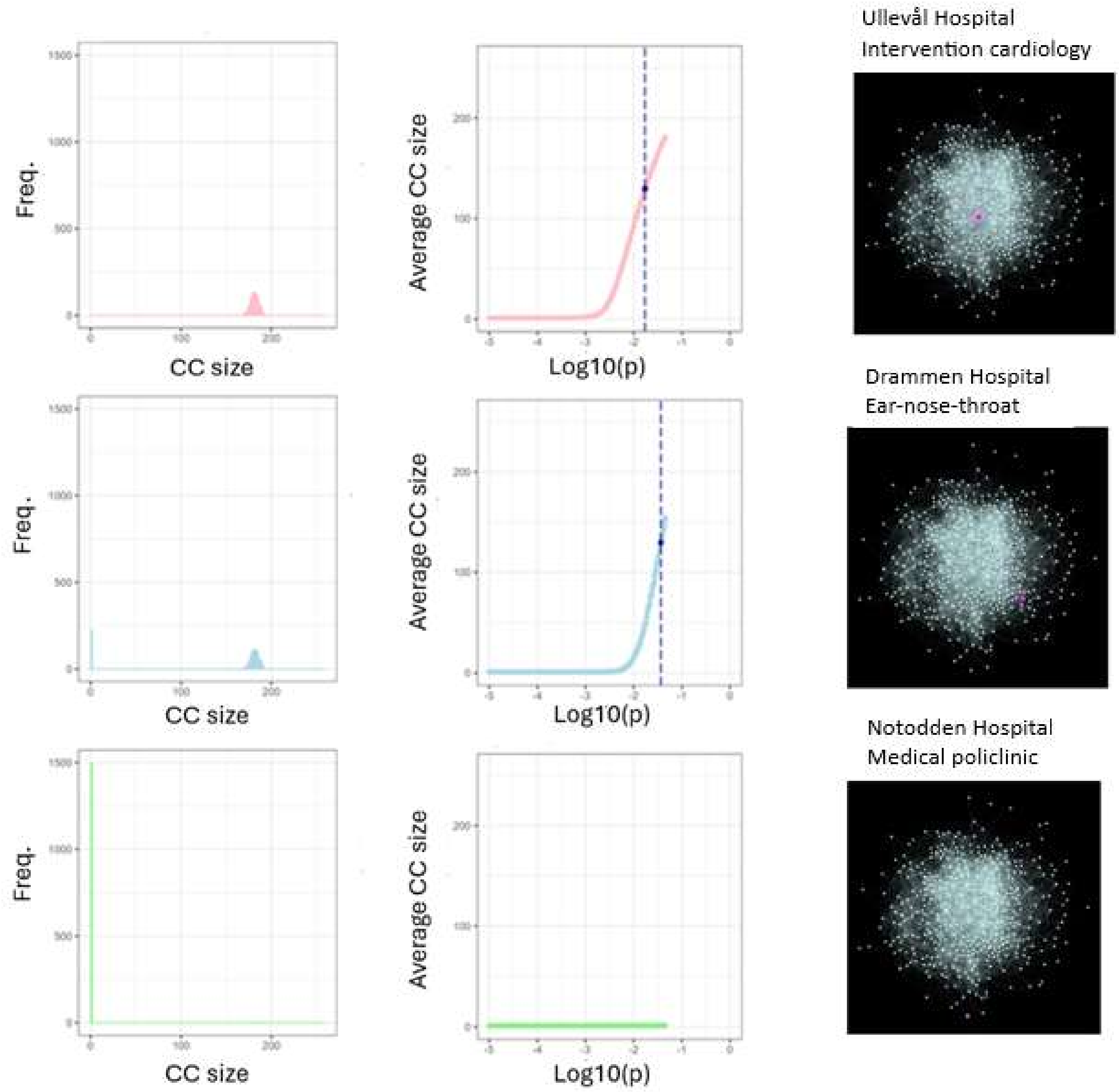


**Fig SuppA.2: Bi-modal percolation behaviour of ward connectivity.** *For three representative wards in the inpatient network (central hub: Ull-IntCard; intermediary: DRA-ENT; fringe ward: NOT-MedPoli), left panels show the distribution of connected component (CC) sizes across 1500 simulations at a fixed edge-retention probability* $p = 10^{-2.2} \sim 6 \cdot 10^{-3}$. *Middle panels show the corresponding average CC sizes as a function of* $\log_{10}(p)$. *Right panels show the position of each ward in the network. As p increases, the CC size distribution becomes bimodal: simulations either result in the ward remaining in a small, isolated component or attaching to the giant connected component. Central wards attach to the giant component at lower values of p, yielding earlier increases in average CC size, while fringe wards remain isolated across most of the parameter range.*

#### B. SI-model simulation of ward-to-ward diffusion and impact of patient movement mediated interventions

We used a simple discrete-time stochastic Susceptible-Infected (SI) model on the directed patient movement networks to examine how the network structure influences diffusion under simplified transmission assumptions. The model is intentionally idealised and is not intended to represent the transmission dynamics of any specific pathogen. Rather, it provides a common framework for ranking wards by their dynamic reachability and exposure and for comparing network-wide diffusion properties of alternative network perturbation scenarios. These perturbations may be interpreted as generic intervention scenarios modifying patient movement within this abstract setting.

**Network:** Each ward is represented as a node in a directed weighted network, where edge weights represent annual frequencies of patient movements derived from either the all-patient or inpatient data. Simulations were performed on the largest weakly connected component (LWCC) of the baseline network. The same set of wards was retained across all intervention scenarios to ensure comparability.

**Diffusion dynamics**: Diffusion was initiated from a single seed ward and followed a discrete-time stochastic Susceptible–Infected (SI) process without recovery. Transmission follows a time-discrete Poisson process approximation in which all outgoing connections from an infected ward carry an infection risk. At each time step, $t$, an infected ward, $w_i$, is assumed to transmit infection to a susceptible ward, $w_j$, with probability

$$p_{ij} = 1 - \exp(-\lambda_{ij}); \qquad \lambda_{ij} = M_{ij}/\alpha$$

Here $M_{ij}$ is the $416 \times 416$ time-aggregated patient movements matrix, and $\alpha$ is an arbitrary scaling factor applied to annual counts ($\alpha = 366$).

Simulation time is an abstract measure of update steps rather than calendar time. Consequently, $p_{ij}$ represents a relative transmission opportunity rather than an absolute infection probability.

**Simulation setup:** Each simulation starts with seeding in a single ward, $w_s$ , and proceeds on the fixed network. The simulation continues up to a maximum horizon of $T_{max} = 500$ time steps. Infection states in the wards are updated across all nodes at each time step.

For each $w_s$, the outward hitting time $H^r_{s,j}$ is the first time step when infection reaches ward $j$ in iteration $r$. If ward $j$ is not reached before $T_{max}$, $w_j$ is assigned a penalised hitting time $H^r_{s,j} = T_{max}$, thereby ensuring comparable summary statistics across scenarios. Each seed ward was simulated in $R = 1000$ independent iterations.

**Computation of summary hitting times and scenario comparison:**

For each seed ward $s$, the *outward mean hitting time* quantifies how long, on average, it takes infection originating in ward $s$ to reach all other wards:

$$\mu^K_{out}(s) = \frac{1}{(N-1)\,R} \sum_{\substack{j=1 \\ j \neq s}}^{N} \sum_{r=1}^{R} \max(H^{r,K}_{s,j}, T_{max})$$

where $K$ denotes the intervention scenario.

The s*cenario-level outward mean hitting time* $\bar{T}^K_{out}$, describing the system-wide diffusion speed, is calculated as a mean over all seeds:

$$\bar{T}_{out}^{K} = \frac{1}{N}\sum_{s=1}^{N} \mu_{out}^{K}(s)$$

Because the model uses an arbitrary scaling constant ($\alpha$) and a fixed censoring horizon, absolute hitting times have no direct temporal interpretation. We therefore compared scenarios using the relative change in network-wide outward mean hitting time:

$$\Delta K(\%) = 100 \times \frac{\bar{T}_{out}^{K} - \bar{T}_{out}^{baseline}}{\bar{T}_{out}^{baseline}}$$

A positive $\Delta K$ indicates a slower diffusion reflecting both reduced network-level connectivity (more censored targets) and longer delays along reachable infection paths. Because we use the same node set, same censoring threshold, differences in $\Delta K$ provide a directly comparable measure on the effects of network perturbation on system-wide diffusion.

**Network perturbation scenarios:** We use as a baseline scenario the unmodified full-year ward-to-ward patient movement network (all-patient or inpatients), which is compared against:

*Edge perturbations*

*Targeted edge removals*: the edges with the highest number of patient movement counts are removed sequentially until a given percentage of flow is eliminated

*Proportional flow reduction*: patient flow on all edges is reduced proportionally by a fixed percentage.

For both, we target flow reductions of 2.5% - 97.5%. Note that for targeted removals, the actual flow reduction may differ slightly from the target for small percentages since the specific edges may carry high volume.

*Node perturbations*
Targeted node interventions were designed to remove wards at highest dynamic exposure. Specifically, we ranked wards by their *inward mean hitting time* in the baseline network, defined as the average time required for infection seeded elsewhere in the network to reach a given ward:

$$\mu_{in}^{basel}(j) = \frac{1}{(N-1)\,R}\sum_{\substack{s=1 \\ s\neq j}}^{N}\sum_{r=1}^{R} \max(H_{s,j}^{r,basel}, T_{max})$$

Wards with the smallest inward mean hitting times (top central) are those that are reached most rapidly on average and are therefore most structurally exposed to incoming patient flow.

*Targeted node removal*: removing *x* (top central) wards with the smallest inward mean hitting time in the baseline.

*Random node removal*: removing *x* wards selected uniformly at random.

We studied moderate node-removal interventions, with the number of removed wards ranging from $x = 5$ to $x = 100$ in the inpatient network and from $x = 5$ to $x = 150$ in the all-patient network. The node removal was implemented by setting its incoming and outgoing patient flows to 0. Removed wards were excluded as seeds but remained as targets, where they receive a penalised hitting time of $T_{max}$.

**Relating dynamic diffusion risk to static network structure:**

To relate the diffusion-based exposure risk to the static network structure, we compared the ward-specific inward mean hitting times with their topological roles in the GMM-derived inflow K-core in the main manuscript.

**Results**

**Consistency between dynamic diffusion and structural backbone**

Wards ranked by minimal global inward mean hitting time in the SI simulations largely coincided with the most central wards identified in the inflow K-core GMM analysis (Table SuppB.1). This close agreement demonstrates that diffusion under the simplified SI process is strongly constrained by the structural backbone identified in the static network analysis.

In the inpatient network, several wards with relatively low patient throughput nevertheless ranked highly by inward hitting time, reflecting strong inbound connectivity from core-central hubs rather than high local flow volumes. In addition, a small number of fast-reached wards outside the central K-core were identified (e.g. *ARE-med* and *AHU-Sten-rehab*), indicating the presence of local feeder structures that are tightly linked to core-central wards despite not belonging to the core themselves.

**Table Supp B1: Top-ranked 20 wards (smallest inward mean hitting times) from the SI simulation model and their corresponding results of the inflow GMM central K-cores.**

| ALL PATIENTS | | | INPATIENTS | | |
|---|---|---|---|---|---|
| Rank | Ward | Inflow GMM centr.K-core | Rank | Ward | Inflow GMM centr. K-core |
| 1 | ULL-intervcard | central hub | 1 | ULL-intervcard | central hub |
| 2 | AHU-heartmed | central hub | 2 | ULL-heart | central hub |
| 3 | ULL-heart | central hub | 3 | AHU-heartmed | central hub |
| 4 | ØST-med | central hub | 4 | ØST-med | central hub |
| 5 | TØN-med | central hub | 5 | ULL-thorax | central intermediary |
| 6 | ULL-thorax | central hub | 6 | TØN-med | central hub |
| 7 | DRA-med | central hub | 7 | DRA-med | central intermediary |
| 8 | ULL- 67 | central hub | 8 | BÆR-med | central intermediary |
| 9 | BÆR-med | central hub | 9 | LOV-med | central intermediary |
| 10 | LOV-med | central hub | 10 | SKI-med | central intermediary |
| 11 | SKI-med | central hub | 11 | DIA-med | central intermediary |
| 12 | RAD-breastonc | central hub | 12 | RIN-med | low-flow stable receiver |
| 13 | DIA-med | central hub | 13 | ARE-med | central intermediary |
| 14 | AHU-neuro | central hub | 14 | AHU-Sten-rehab | - |
| 15 | RAD-lungonc | central hub | 15 | ULL-acutemed | central intermediary |
| 16 | RAD-uroonc | central hub | 16 | KRI-med | central intermediary |
| 17 | AHU-ort | central hub | 17 | ØST-surg | low-flow stable receiver |
| 18 | TØN-surg | central hub | 18 | GJØ-intmed | low-flow stable receiver |
| 19 | ARE-med | - | 19 | RIK-neuroF2 | central intermediary |
| 20 | RIN-med | central intermediary | 20 | RIK-lungmed | central intermediary |

##### Edge-based perturbations

Across both networks, uniform reduction of patient flow consistently produced larger increases in mean hitting time (ΔK) than targeted removal of high-flow edges at low to intermediate levels of patient flow reductions (Fig. SuppB.1). These findings suggest that slowing large-scale diffusion is achieved more effectively by weakening many transmission pathways than by eliminating a small number of high-flow connections.

The magnitude of ΔK was systematically smaller in the inpatient network, consistent with its smaller size and more compact, cohesive structure. In both networks, targeted edge removal had little impact on ΔK up to approximately 50% flow reduction, suggesting substantial redundancy in routing through the dense network core. Only close to network collapse—at flow reductions exceeding 90–95%—did targeted removal become comparatively effective, as residual transmission became concentrated in a small number of critical edges.

Overall, these results indicate that distributed reductions in patient movement are more disruptive to network-wide diffusion than selective removal of high-flow connections, except in the near-critical connectivity regime. Larger and more redundant networks therefore require more extensive perturbations to achieve structural fragmentation.

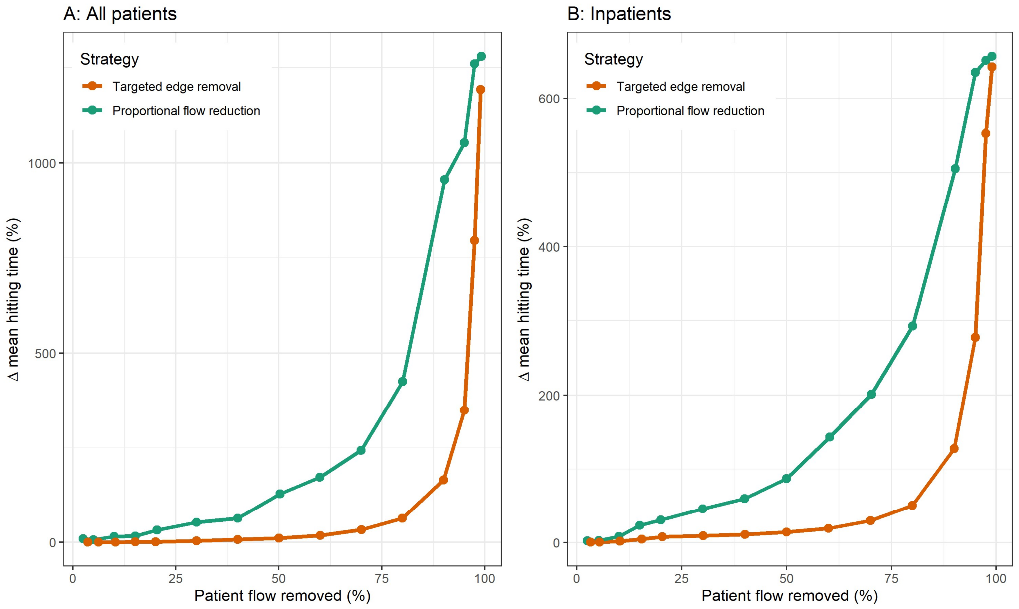


*Fig SuppB.1: Changes in mean hitting time ΔK (%) under uniform and targeted edge-based removal for the all-patient and inpatient networks.*

##### Node-based perturbations

Removing wards substantially delayed network-wide diffusion in both networks, with targeted removal consistently outperforming random removal and producing larger increases in mean hitting time (ΔK) for the same number of wards removed (Table SuppB.2; Fig. SuppB.2). This advantage was accompanied by markedly greater reductions in total patient flow, indicating that structurally embedded wards concentrate both connectivity and movement volume.

**Table SuppB.2: Changes in mean hitting time ΔK (%) under uniform and targeted node-based interventions for all-patient and inpatient networks.**

| | **All-patient network** | | | | **Inpatient network** | | | |
|---|---|---|---|---|---|---|---|---|
| | Random node removal | | Targeted node removal | | Random node removal | | Targeted node removal | |
| Nodes removed | ΔK(%) | Flow red. | ΔK(%) | Flow red. | ΔK(%) | Flow red. | ΔK(%) | Flow red. |
| 5 | 16,5 | 1,9 | 20,6 | 15,5 | 12,8 | 2 | 21,3 | 26,8 |
| 10 | 36 | 5,2 | 43,7 | 32,7 | 29,4 | 8,3 | 42,4 | 38 |
| 15 | 49,4 | 8,6 | 66,7 | 38,6 | 39,1 | 9,4 | 65 | 47,7 |
| 20 | 68,5 | 14,8 | 91 | 44,7 | 59,9 | 20,6 | 87,3 | 55,1 |
| 30 | 96,6 | 11,8 | 136 | 55,3 | 81,6 | 12,7 | 127,2 | 64,4 |
| 40 | 134,9 | 14,4 | 176,5 | 63,6 | 118,7 | 26,3 | 158,5 | 69 |
| 50 | 168,5 | 27,3 | 213,9 | 68 | 143,4 | 42 | 196,8 | 74,3 |
| 60 | 204 | 26,1 | 252,2 | 71,1 | 155,2 | 25,3 | 236,5 | 77,7 |
| 80 | 263,8 | 37,2 | 340,7 | 77,6 | 216,9 | 52,8 | 318,3 | 86,8 |
| 100 | 336,3 | 44,7 | 415,7 | 82,8 | 270,3 | 55,7 | 374,9 | 92,1 |
| 125 | 422,4 | 43,4 | 530,7 | 87,7 | | | | |
| 150 | 499,0 | 60,9 | 630,0 | 90,9 | | | | |

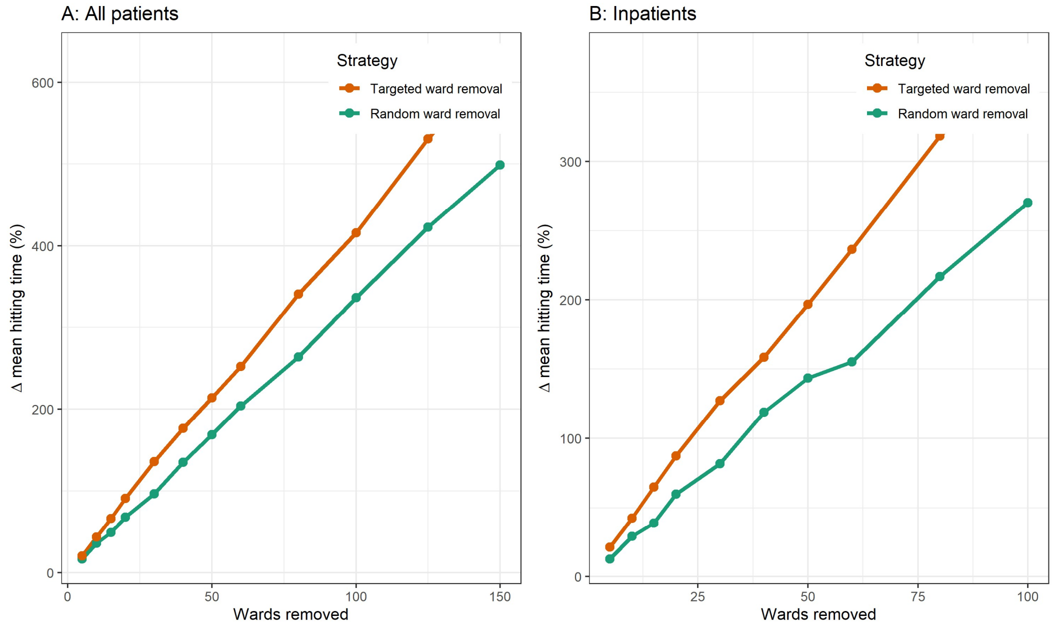


*Fig SuppB.2: Changes in mean hitting time ΔK (%) under random and targeted ward removal in the all-patient and inpatient networks.*

As in the edge-based interventions, the inpatient network exhibited smaller ΔK values than the all-patient network, reflecting its more compact structure and lower overall redundancy. Node (ward) removal simultaneously alters network topology, reduces total patient flow, and constrains the set of possible seeds. The resulting increases in hitting time therefore reflect a combination of structural fragmentation, reduced movement and reduced seeding opportunities, and are not directly comparable with the edge-based perturbation scenarios.

#### C. Illustrative application to the 2012 Østfold Hospital VRE outbreak

While the previous section focused on system-level diffusion and the effects of patient movement–based interventions across the entire hospital network, outbreak investigations typically begin with transmission originating from a specific ward. To illustrate how the network-based framework can be applied at this local scale, we retrospectively analysed a well-characterised outbreak of vancomycin-resistant *Enterococcus* (VRE) by linking individual-level patient admission records with notified VRE cases from the Norwegian Surveillance System for Communicable Diseases (MSIS).

In 2012, Østfold Hospital experienced a large outbreak of VRE (vanA) that extended into 2013. Subsequent epidemiological investigations suggested that transmission began silently in June 2012 and remained undetected until August, when identification of a VRE-positive patient in the surgical ward triggered extensive screening (4). The outbreak primarily affected patients admitted to the gastric surgery ward (*ØST-surg*) and the infectious diseases and pulmonary wards (*ØST-med*), with a smaller number of cases identified in the orthopaedic ward (*ØST-ort*). Because the patient administrative system at Østfold Hospital comprised relatively few organisational units during the study period, each registered ward typically represented several clinical specialities.

A total of 79 VRE-positive cases (68 *vanA*) were reported to MSIS during 2012. Restricting the analysis to *vanA*-positive patients reported between 1 June and 31 December 2012 who had been admitted to Østfold Hospital on or before the date of their first positive test yielded 66 eligible cases. Of these, 61 patients were hospitalised at Østfold Hospital when VRE was detected. The remaining vanA cases included two patients who tested positive in Oslo-based hospitals (Rikshospitalet and Ullevål; wards not specified due to privacy constraints and low case counts), while a few patients were not hospitalised at the time of testing.

Using ØST-surg as the presumed outbreak source, the network analysis ranked ØST-med and ØST-ort among the wards with the shortest diffusion paths and strong structural connectivity to the index ward (Table Supp C.2). These were also the principal wards in which cases were identified during the outbreak, illustrating close correspondence between the network-derived rankings and the observed spatial distribution of cases. In contrast, the two patients detected in Oslo-based hospitals were not associated with wards that ranked highly in terms of network reachability from ØST-surg. This discrepancy may partly reflect the relatively coarse ward classification available for Østfold Hospital during the study period, which limited the resolution of the patient movements and may have obscured specific referral pathways.

Overall, the outbreak appeared largely confined to Østfold Hospital, suggesting that locally available administrative data on internal patient movements can be useful for guiding and prioritising investigation and screening efforts. This retrospective case study demonstrates the practical value of patient movement networks for identifying wards most closely connected to a potential outbreak source, while also highlighting that movement data should be interpreted alongside pathogen-specific epidemiological information, microbiological evidence, and sufficiently detailed organisational data.

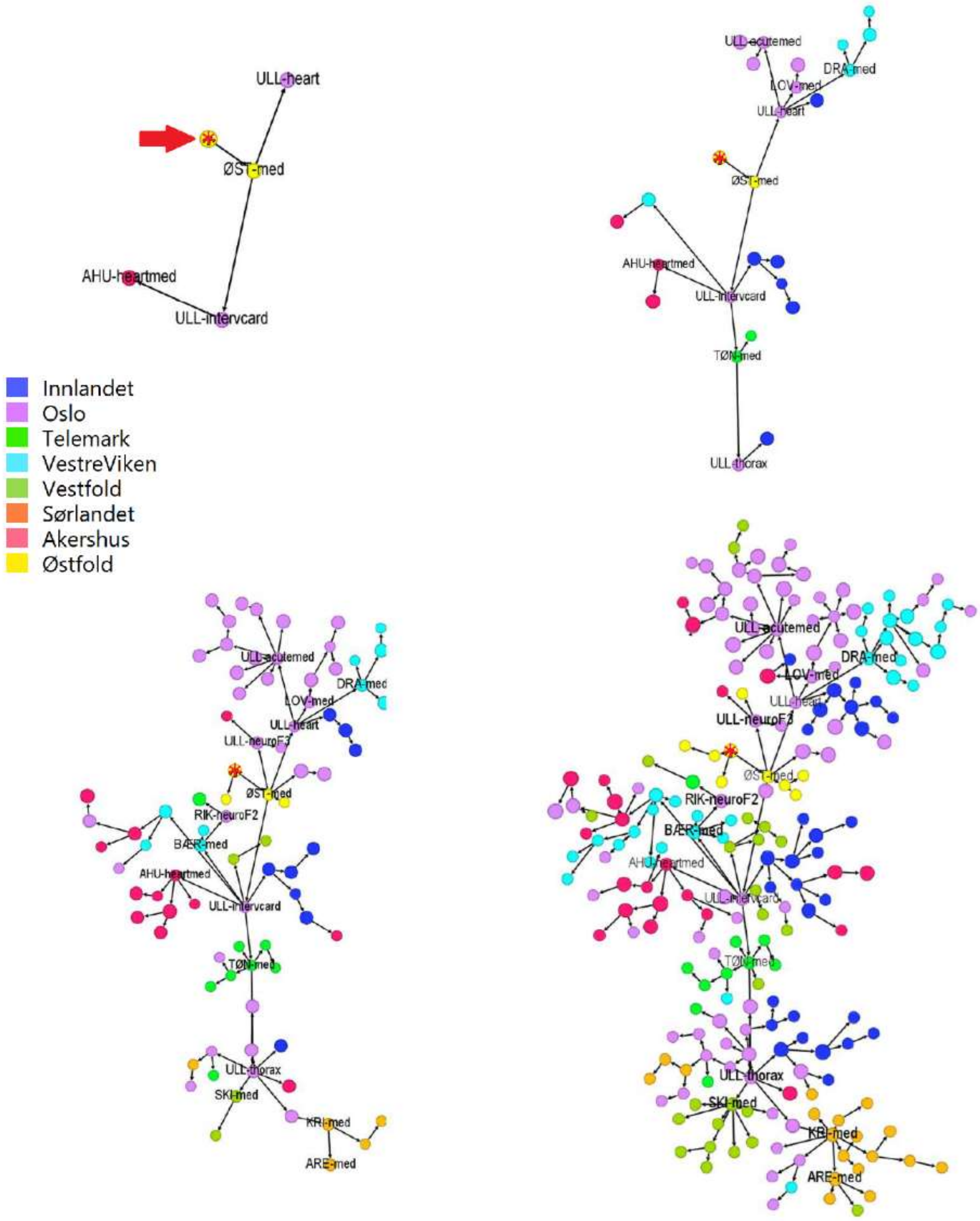


*Fig. Supp C.1: Example realisation of network growth using the SI model seeded in the surgical ward at Østfold Hospital (indicated by an arrow and*), illustrating the temporal expansion of the network using inpatient patient-movement data. Wards are colour-coded by region, with selected larger wards labelled.*

***Table Supp C.2: Top 25 wards closest to the surgical ward at Østfold Hospital based on patient-flow diffusion simulations.*** *Wards are ranked separately in the all-patient network and the inpatient-only network by their outward mean penalised hitting time from the index ward (ward 298), estimated from 1,000 stochastic SI simulations with a simulation horizon of T=500 time steps. Lower values indicate faster dynamic reachability from the surgical ward to other wards. A combined score is also reported, defined as the sum of percentile ranks across both networks, identifying wards that are consistently close to the index ward in both networks.*

| | **Inpatient network** | | **All-patient network** | | **Combined score** | |
|---|---|---|---|---|---|---|
| **Rank** | **ward** | **mean** | **ward** | **mean** | **ward** | **score** |
| 1 | ØST-med | 1,128 | ØST-med | 1,502 | ØST-med | 0,006 |
| 2 | ØST-ort | 2,361 | ULL-intervcard | 3,113 | ULL-intervcard | 0,019 |
| 3 | ØST-phys | 2,379 | ULL-heart | 3,377 | ØST-ort | 0,020 |
| 4 | ØST-women | 2,394 | ØST-ort | 3,601 | ULL-heart | 0,030 |
| 5 | ULL-intervcard | 2,686 | AHU-heartmed | 3,858 | ØST-neuro | 0,037 |
| 6 | ØST-neuro | 2,708 | ØST-neuro | 3,92 | AHU-heartmed | 0,045 |
| 7 | ØST-ent | 2,883 | ULL-thorax | 4,165 | ØST-women | 0,054 |
| 8 | ULL-heart | 2,943 | TØN-med | 4,178 | ULL-thorax | 0,055 |
| 9 | ØST-eye | 3,001 | DRA-med | 4,315 | TØN-med | 0,061 |
| 10 | ØST-ped | 3,385 | BÆR-med | 4,513 | DRA-med | 0,070 |
| 11 | AHU-heartmed | 3,47 | LOV-med | 4,78 | BÆR-med | 0,078 |
| 12 | ULL-thorax | 3,715 | ØST-women | 4,798 | LOV-med | 0,087 |
| 13 | TØN-med | 3,782 | RIN-med | 4,822 | RIN-med | 0,099 |
| 14 | ØST-rheu | 3,792 | DIA-med | 5,021 | DIA-med | 0,100 |
| 15 | DRA-med | 3,864 | ULL-acutemed | 5,039 | ULL-acutemed | 0,114 |
| 16 | ULL- 67 | 3,886 | GJØ-intmed | 5,246 | ULL-neuroF3 | 0,126 |
| 17 | BÆR-med | 4,035 | ULL-neuroF3 | 5,304 | GJØ-intmed | 0,127 |
| 18 | RAD-breastonc | 4,112 | LIL-intmed | 5,46 | RIK-lungmed | 0,131 |
| 19 | LOV-med | 4,222 | RIK-lungmed | 5,488 | ØST-ped | 0,135 |
| 20 | DIA-med | 4,343 | AHU-Sten-rehab | 5,53 | LIL-intmed | 0,142 |
| 21 | RIN-med | 4,358 | HAMELV-intmed | 5,616 | RAD-breastonc | 0,151 |
| 22 | RAD-lungonc | 4,366 | KOB-med | 5,652 | AHU-Sten-rehab | 0,156 |
| 23 | RAD-uroonc | 4,373 | LAR-med | 5,657 | HAMELV-intmed | 0,165 |
| 24 | ULL-acutemed | 4,385 | RIK-neuroF2 | 5,714 | RIK-neuroF2 | 0,169 |
| 25 | RIK-lungmed | 4,489 | HAM-intmed | 5,807 | ØST-rheu | 0,171 |

### Supplementary tables

##### Table S1: Descriptive statistics of hospitals in South-Eastern Health Region (2012)

| | | | Registrations | | Prevalence | | Treatment (%) | | |
|---|---|---|---|---|---|---|---|---|---|
| **Name** | **region** | **wards** | **count** | **% (total)** | **daily mean** | **% (total)** | **Inpat** | **Polyclin.** | **Day pat.** |
| All | | 416 | 3618299 | 100,0 | 15109 | 100,0 | 14,6 | 78,9 | 6,5 |
| Akershus | Akershus | 25 | 365611 | 10,1 | 1648 | 10,9 | 19,0 | 74,0 | 7,0 |
| Sunnaas | | 3 | 6075 | 0,2 | 125 | 0,8 | 45,6 | 54,2 | 0,2 |
| LillehammerRheu | Innlandet | 1 | 15332 | 0,4 | 69 | 0,5 | 9,2 | 0,0 | 90,8 |
| HamarElverum | | 9 | 163168 | 4,5 | 650 | 4,3 | 14,4 | 77,1 | 8,4 |
| Gjøvik | | 7 | 96340 | 2,7 | 410 | 2,7 | 14,5 | 79,6 | 6,0 |
| Granheim | | 1 | 2031 | 0,1 | 38 | 0,3 | 24,3 | 0,0 | 75,7 |
| Kongsvinger | | 9 | 55171 | 1,5 | 230 | 1,5 | 14,5 | 82,4 | 3,1 |
| Lillehammer | | 6 | 95431 | 2,6 | 420 | 2,8 | 18,3 | 73,6 | 8,1 |
| Ottestad | | 2 | 11238 | 0,3 | 54 | 0,4 | 7,0 | 93,0 | 0,0 |
| Tynset | | 3 | 14760 | 0,4 | 71 | 0,5 | 23,0 | 72,3 | 4,7 |
| Diakonhjemmet | Oslo | 4 | 80923 | 2,2 | 337 | 2,2 | 13,6 | 84,1 | 2,3 |
| Lovisenberg | | 6 | 71773 | 2,0 | 308 | 2,0 | 15,1 | 76,9 | 8,0 |
| OlafiaClinic | | 1 | 25182 | 0,7 | 69 | 0,5 | 2,2 | 0,0 | 97,8 |
| OsloPubEmerg (Oslo Legevakt) | | 1 | 7457 | 0,2 | 32 | 0,2 | 100,0 | 0,0 | 0,0 |
| Aker | | 15 | 74563 | 2,1 | 274 | 1,8 | 9,3 | 82,2 | 8,5 |
| Radiumhospitalet | | 11 | 95899 | 2,7 | 409 | 2,7 | 9,4 | 90,2 | 0,4 |
| Rikshospitalet | | 42 | 203192 | 5,6 | 962 | 6,4 | 16,2 | 74,7 | 9,1 |
| Ullevål | | 67 | 602132 | 16,6 | 2287 | 15,1 | 11,2 | 82,9 | 5,9 |
| Arendal | Sørlandet | 12 | 128711 | 3,6 | 500 | 3,3 | 13,7 | 78,7 | 7,6 |
| Flekkefjord | | 5 | 28656 | 0,8 | 125 | 0,8 | 18,0 | 73,2 | 8,9 |
| Kristiansand | | 16 | 215801 | 6,0 | 841 | 5,6 | 13,0 | 82,6 | 4,4 |
| Betanien | Telemark | 9 | 24214 | 0,7 | 86 | 0,6 | 7,8 | 82,5 | 9,7 |
| Kragerø | | 9 | 13017 | 0,4 | 69 | 0,5 | 13,0 | 79,4 | 7,6 |
| Notodden | | 16 | 20850 | 0,6 | 91 | 0,6 | 22,0 | 72,5 | 5,5 |
| Rjukan | | 14 | 7566 | 0,2 | 41 | 0,3 | 25,1 | 67,0 | 7,9 |
| SkienPorsgrunn | | 39 | 184475 | 5,1 | 755 | 5,0 | 14,8 | 79,3 | 5,9 |
| Larvik | Vestfold | 3 | 21612 | 0,6 | 73 | 0,5 | 5,7 | 83,6 | 10,7 |
| Vestfold | | 5 | 20463 | 0,6 | 96 | 0,6 | 10,7 | 58,4 | 30,9 |
| TønsbergHab | | 1 | 3128 | 0,1 | 9 | 0,1 | 100,0 | 0,0 | 0,0 |
| Tønsberg | | 10 | 235137 | 6,5 | 955 | 6,3 | 15,0 | 79,9 | 5,1 |
| Bærum | VestreViken | 9 | 106566 | 2,9 | 480 | 3,2 | 17,6 | 74,6 | 7,7 |
| Drammen | | 18 | 213595 | 5,9 | 888 | 5,9 | 14,4 | 79,4 | 6,2 |
| Kongsberg | | 9 | 33994 | 0,9 | 152 | 1,0 | 19,7 | 71,4 | 8,9 |
| MartinaHansen | | 5 | 35198 | 1,0 | 135 | 0,9 | 8,4 | 84,2 | 7,4 |
| Ringerike | | 13 | 60883 | 1,7 | 274 | 1,8 | 19,5 | 71,1 | 9,4 |
| Østfold | Østfold | 12 | 278155 | 7,7 | 1145 | 7,6 | 16,1 | 77,3 | 6,6 |

**Table S2: Patient-level distributions of registration, episode and inter-ward movement counts with cumulative proportions: all patients (top, N=984,437) and inpatients (bottom; N=322,593).** The proportion of patients registered as residents of HSØ was 96.4% and 95.9% for all patients, and inpatients, respectively.

**All patients (N=984 437)**

| | Registrations (range 1-377) | | | | Episodes (range 1-258) | | | | Movements (range 0-198) | | | |
|---|---|---|---|---|---|---|---|---|---|---|---|---|
| Value | Pat. No. | Cum. Prop Pat. (%) | Reg. No. | Cum. Prop Reg. (%) | Pat. No. | Cum. Prop Pat.(%) | Epi. No. | Cum. Prop Epi. (%) | Pat. No. | Cum. Prop Pat. (%) | Mov. No. | Cum. Prop Mov. (%) |
| 0 | | | | | | | | | 898 417 | 91,3 | | |
| 1 | 388 489 | 39,5 | 388 489 | 10,7 | 408 302 | 41,5 | 408 302 | 12,3 | 54 377 | 96,8 | 54 377 | 34,0 |
| 2 | 195 455 | 59,3 | 390 910 | 21,5 | 197 237 | 61,5 | 394 474 | 24,2 | 17 377 | 98,6 | 34 754 | 55,7 |
| 3 | 118 015 | 71,3 | 354 045 | 31,3 | 118 040 | 73,5 | 354 120 | 34,8 | 6 177 | 99,2 | 18 531 | 67,2 |
| 4 | 76 272 | 79,1 | 305 088 | 39,8 | 74 928 | 81,1 | 299 712 | 43,8 | 2 907 | 99,5 | 11 628 | 74,5 |
| 5 | 50 623 | 84,2 | 253 115 | 46,8 | 48 555 | 86,0 | 242 775 | 51,1 | 1 651 | 99,6 | 8 255 | 79,6 |
| 6 | 34 667 | 87,7 | 208 002 | 52,5 | 32 507 | 89,3 | 195 042 | 57,0 | 1 013 | 99,7 | 6 078 | 83,4 |
| 7 | 24 650 | 90,2 | 172 550 | 57,3 | 22 706 | 91,7 | 158 942 | 61,8 | 659 | 99,8 | 4 613 | 86,3 |
| 8 | 17 831 | 92,0 | 142 648 | 61,2 | 16 021 | 93,3 | 128 168 | 65,7 | 430 | 99,9 | 3 440 | 88,5 |
| 9 | 13 298 | 93,4 | 119 682 | 64,5 | 11 755 | 94,5 | 105 795 | 68,8 | 315 | 99,9 | 2 835 | 90,2 |
| 10 | 10 185 | 94,4 | 101 850 | 67,3 | 8 933 | 95,4 | 89 330 | 71,5 | 257 | 99,9 | 2 570 | 91,8 |
| 11-20 | 37 891 | 98,3 | 532 246 | 82,0 | 31 876 | 98,6 | 445 072 | 84,9 | 770 | 100,0 | 10 292 | 98,3 |
| 20+ | 17 061 | 100,0 | 649 674 | 100,0 | 13 577 | 100,0 | 501 095 | 100,0 | 87 | 100,0 | 2 780 | 100,0 |

**Inpatients (N=322 593)**

| | Registrations (range 1-138) | | | | Episodes (range 1-50) | | | | Movements (range 0-112) | | | |
|---|---|---|---|---|---|---|---|---|---|---|---|---|
| Value | Pat. No. | Cum. Prop Pat. (%) | Reg. No. | Cum. Prop Reg. (%) | Pat. No. | Cum. Prop Pat. (%) | Epi. No. | Cum. Prop Epi. (%) | Pat. No. | Cum. Prop Pat. (%) | Mov. No. | Cum. Prop Mov. (%) |
| 0 | | | | | | | | | 285 130 | 88,4 | | |
| 1 | 224 690 | 69,7 | 224 690 | 42,4 | 245 056 | 76,0 | 245 056 | 52,8 | 23 824 | 95,8 | 23 824 | 39,3 |
| 2 | 53 248 | 86,2 | 106 496 | 62,5 | 47 891 | 90,8 | 95 782 | 73,5 | 9 076 | 98,6 | 18 152 | 69,3 |
| 3 | 21 359 | 92,8 | 64 077 | 74,6 | 15 524 | 95,6 | 46 572 | 83,5 | 2 505 | 99,4 | 7 515 | 81,7 |
| 4 | 9 686 | 95,8 | 38 744 | 81,9 | 6 704 | 97,7 | 26 816 | 89,3 | 1 079 | 99,7 | 4 316 | 88,8 |
| 5 | 5 066 | 97,4 | 25 330 | 86,7 | 3 128 | 98,7 | 15 640 | 92,7 | 428 | 99,8 | 2 140 | 92,3 |
| 6 | 3 031 | 98,3 | 18 186 | 90,2 | 1 674 | 99,2 | 10 044 | 94,9 | 235 | 99,9 | 1 410 | 94,7 |
| 7 | 1 833 | 98,9 | 12 831 | 92,6 | 976 | 99,5 | 6 832 | 96,3 | 111 | 99,9 | 777 | 95,9 |
| 8 | 1 181 | 99,2 | 9 448 | 94,4 | 577 | 99,7 | 4 616 | 97,3 | 74 | 100,0 | 592 | 96,9 |
| 9 | 787 | 99,5 | 7 083 | 95,7 | 348 | 99,8 | 3 132 | 98,0 | 40 | 100,0 | 360 | 97,5 |
| 10 | 463 | 99,6 | 4 630 | 96,6 | 201 | 99,8 | 2 010 | 98,4 | 24 | 100,0 | 240 | 97,9 |
| 11-20 | 1 153 | 100,0 | 15 334 | 99,5 | 475 | 100,0 | 6 262 | 99,8 | 52 | 100,0 | 700 | 99,1 |
| 20+ | 96 | 100,0 | 2 805 | 100,0 | 39 | 100,0 | 1 033 | 100,0 | 15 | 100,0 | 564 | 100,0 |

**Table S3: Inter and intra-regional patient ward transfers during the study period for all patients (top) and inpatients only (bottom).** Each cell reports the share (%) of all transfers from the *row region* that were directed to the *column region* (row percentages sum to 100%). Diagonal cells showing intra-region transfers are highlighted in blue.

All patients (N=160,153)

| | Akershus | Innlandet | Oslo | Sørlandet | Telemark | Vestfold | VestreViken | Østfold |
|---|---|---|---|---|---|---|---|---|
| Akershus | 85,2 | 0,4 | 13,4 | 0 | 0,1 | 0,1 | 0,5 | 0,3 |
| Innlandet | 1,2 | 85,7 | 12,4 | 0 | 0 | 0,1 | 0,3 | 0,2 |
| Oslo | 5,7 | 2,8 | 80 | 1,3 | 1,1 | 2,1 | 4,5 | 2,5 |
| Sørlandet | 0,1 | 0 | 4 | 92,8 | 3 | 0,1 | 0,1 | 0 |
| Telemark | 0,2 | 0 | 5,3 | 3,5 | 88,9 | 1,7 | 0,3 | 0 |
| Vestfold | 0,2 | 0,1 | 12,5 | 0,2 | 2,4 | 83,4 | 1,1 | 0,1 |
| VestreViken | 0,7 | 0,1 | 14,7 | 0 | 0,2 | 0,5 | 83,7 | 0,1 |
| Østfold | 1,3 | 0 | 13,9 | 0,1 | 0 | 0,2 | 0,2 | 84,3 |

Inpatients (N=60,590)

| | Akershus | Innlandet | Oslo | Sørlandet | Telemark | Vestfold | VestreViken | Østfold |
|---|---|---|---|---|---|---|---|---|
| Akershus | 75,3 | 0,9 | 22,4 | 0 | 0,1 | 0,1 | 0,6 | 0,6 |
| Innlandet | 2,3 | 72 | 24,5 | 0,1 | 0,1 | 0,1 | 0,6 | 0,3 |
| Oslo | 10,4 | 6,6 | 57 | 3,2 | 2,6 | 5 | 9,6 | 5,6 |
| Sørlandet | 0,2 | 0 | 9,2 | 82,5 | 7,7 | 0,2 | 0,2 | 0 |
| Telemark | 0,3 | 0,1 | 11,7 | 8,3 | 76,2 | 2,8 | 0,5 | 0 |
| Vestfold | 0,4 | 0,3 | 28 | 0,4 | 3 | 66 | 1,8 | 0,1 |
| VestreViken | 1 | 0,2 | 29,1 | 0,1 | 0,3 | 0,8 | 68,3 | 0,3 |
| Østfold | 3 | 0,1 | 31,5 | 0,2 | 0,1 | 0,5 | 0,6 | 64 |

**Table S4: Global network properties by delay in all-patient and inpatient networks**

| Network | Attribute | Baseline 0 days | Lag 14 days | Lag 30 days | Lag 90 days |
|---|---|---|---|---|---|
| All-patient | count nodes | 416 | 416 | 416 | 416 |
| | count conn. nodes | 415 | 416 | 416 | 416 |
| | count edges | 160 153 | 472 357 | 621 321 | 839 939 |
| | count unique edges | 12 626 | 33 715 | 39 398 | 45 896 |
| | edge density | 0,07 | 0,20 | 0,23 | 0,27 |
| | Reciprocity | 0,73 | 0,81 | 0,83 | 0,85 |
| | mean degree | 60,70 | 162,09 | 189,41 | 220,65 |
| | centr.in-degree | 0,28 | 0,39 | 0,42 | 0,44 |
| | centr out-degree | 0,30 | 0,40 | 0,42 | 0,43 |
| | assortativity strength | 0,30 | 0,30 | 0,24 | 0,15 |
| | assortativity degree | 0,11 | 0,01 | -0,01 | -0,03 |
| | mean av. short distance | 2,39 | 1,88 | 1,82 | 1,77 |
| Inpatient | count nodes | 416 | 416 | 416 | 416 |
| | count conn. nodes | 271 | 275 | 275 | 276 |
| | count edges | 60 590 | 76 458 | 86 035 | 104 219 |
| | count unique edges | 5 834 | 7 593 | 8 593 | 10 416 |
| | edge density | 0,03 | 0,04 | 0,05 | 0,06 |
| | Reciprocity | 0,62 | 0,67 | 0,69 | 0,72 |
| | mean degree | 28,05 | 36,50 | 41,31 | 50,08 |
| | centr.in-degree | 0,20 | 0,20 | 0,22 | 0,22 |
| | centr out-degree | 0,20 | 0,21 | 0,21 | 0,22 |
| | assortativity strength | 0,30 | 0,32 | 0,31 | 0,30 |
| | assortativity degree | 0,06 | 0,07 | 0,07 | 0,06 |
| | mean av. short distance | 2,37 | 2,23 | 2,17 | 2,09 |

**Table S5: Persistence of the central core by delay in all-patient and inpatient networks: Baseline retention and jaccard.** The table compares the central K-core at baseline (0 days) with cores identified after 14, 30, and 90 days ¨delay for inflow and outflow networks; $n_{(baseline)}$ and $n_{(lag)}$ denote the number of core wards at baseline and at the given lag, and wards kept from baseline represents their intersection. Retention quantifies baseline persistence, i.e. $|Baseline \cap Lag|/|Baseline|$. The Jaccard index measures overall similarity accounting for both retained and newly appearing wards, $|Baseline \cap Lag|/|Baseline \cup Lag|$. Hence, lower Jaccard values despite high retention indicate core expansion or turnover rather than simple persistence.

| Network | Direction | Lag (days) | N baseline | N lag | Wards kept from baseline | Retention | Jaccard |
|---|---|---|---|---|---|---|---|
| All-patient | Inflow | 14 | 21 | 24 | 18 | 0,86 | 0,67 |
| | | 30 | 21 | 19 | 14 | 0,67 | 0,54 |
| | | 90 | 21 | 19 | 13 | 0,62 | 0,48 |
| | Outflow | 14 | 19 | 13 | 11 | 0,58 | 0,52 |
| | | 30 | 19 | 29 | 14 | 0,74 | 0,41 |
| | | 90 | 19 | 33 | 13 | 0,68 | 0,33 |
| Inpatient | Inflow | 14 | 15 | 12 | 12 | 0,80 | 0,80 |
| | | 30 | 15 | 13 | 13 | 0,87 | 0,87 |
| | | 90 | 15 | 26 | 15 | 1,00 | 0,58 |
| | Outflow | 14 | 11 | 10 | 9 | 0,82 | 0,75 |
| | | 30 | 11 | 32 | 11 | 1,00 | 0,34 |
| | | 90 | 11 | 27 | 11 | 1,00 | 0,41 |

**Table S6: Table S6. Cross-regional mixing by delay.** The table reports the number and share of multi-regional communities (excluding single-ward communities), as well as the wards and hospitals involved in multi-regional patient movements, for all-patient and inpatient networks at baseline (0 days) and delays of 14, 30, and 90 days. *N communities* denotes the total number of multi-ward detected communities; multi-regional communities include wards from more than one region. Reported shares represent the fraction of communities, wards, and hospitals that are multi-regional.

| Network | Delay (days) | Base-line Wards | N Comm. | Multi-reg. Comm. | Share | Wards in Multi-reg. Comm. | Share | Hospitals in Multi-reg. Comm. | Share |
|---|---|---|---|---|---|---|---|---|---|
| All-patient | 0 | 406 | 16 | 5 | 0.312 | 252 | 0.621 | 28 | 0.778 |
| | 14 | 406 | 12 | 4 | 0.333 | 220 | 0.542 | 26 | 0.722 |
| | 30 | 406 | 14 | 4 | 0.286 | 218 | 0.537 | 26 | 0.722 |
| | 90 | 406 | 14 | 3 | 0.214 | 199 | 0.491 | 21 | 0.583 |
| Inpatient | 0 | 264 | 9 | 4 | 0.444 | 224 | 0.848 | 32 | 0.941 |
| | 14 | 264 | 10 | 6 | 0.6 | 229 | 0.867 | 32 | 0.941 |
| | 30 | 264 | 10 | 6 | 0.6 | 215 | 0.814 | 29 | 0.853 |
| | 90 | 264 | 8 | 4 | 0.5 | 179 | 0.681 | 25 | 0.735 |

**Table S7: Strong and weak global percolation thresholds from random edge thinning (1000 simulations) in the all-patient and inpatient networks.** Columns show the mean median and interquartile range of the percolation threshold in probability units (p) at which the largest component reaches $N_w/2$ wards, where $N_w$ is the number of connected wards. For comparison Cp-values were converted to “days” over a 366-day horizon.

| Strong connected component | | | | | | | | | |
|---|---|---|---|---|---|---|---|---|---|
| Network | Delay | | Cp | | | | Cp ("days") | | |
| | (days) | N_w | mean | median | Q25 | Q75 | median | Q25 | Q75 |
| All patients | 0 | 415 | 9,91E-03 | 9,90E-03 | 9,35E-03 | 1,04E-02 | 3,62 | 3,42 | 3,82 |
| | 14 | 416 | 2,34E-03 | 2,33E-03 | 2,21E-03 | 2,45E-03 | 0,85 | 0,81 | 0,90 |
| | 30 | 416 | 1,71E-03 | 1,71E-03 | 1,63E-03 | 1,79E-03 | 0,63 | 0,60 | 0,65 |
| | 90 | 416 | 1,23E-03 | 1,23E-03 | 1,17E-03 | 1,28E-03 | 0,45 | 0,43 | 0,47 |
| Inpatients | 0 | 271 | 1,87E-02 | 1,86E-02 | 1,73E-02 | 2,00E-02 | 6,80 | 6,31 | 7,33 |
| | 14 | 275 | 1,31E-02 | 1,30E-02 | 1,22E-02 | 1,39E-02 | 4,76 | 4,45 | 5,10 |
| | 30 | 275 | 1,10E-02 | 1,10E-02 | 1,03E-02 | 1,17E-02 | 4,02 | 3,78 | 4,28 |
| | 90 | 276 | 8,41E-03 | 8,37E-03 | 7,86E-03 | 8,91E-03 | 3,06 | 2,88 | 3,26 |
| Weak connected component | | | | | | | | | |
| Network | Delay | | Cp | | | | Cp ("days") | | |
| | (days) | N_w | mean | median | Q25 | Q75 | median | Q25 | Q75 |
| All patients | 0 | 415 | 2,96E-03 | 2,94E-03 | 2,78E-03 | 3,15E-03 | 1,08 | 1,02 | 1,15 |
| | 14 | 416 | 7,59E-04 | 7,52E-04 | 7,10E-04 | 8,02E-04 | 0,28 | 0,26 | 0,29 |
| | 30 | 416 | 5,55E-04 | 5,52E-04 | 5,19E-04 | 5,86E-04 | 0,20 | 0,19 | 0,21 |
| | 90 | 416 | 3,99E-04 | 3,98E-04 | 3,75E-04 | 4,20E-04 | 0,15 | 0,14 | 0,15 |
| Inpatients | 0 | 271 | 5,05E-03 | 5,02E-03 | 4,64E-03 | 5,43E-03 | 1,84 | 1,70 | 1,99 |
| | 14 | 275 | 3,75E-03 | 3,74E-03 | 3,45E-03 | 4,03E-03 | 1,37 | 1,26 | 1,47 |
| | 30 | 275 | 3,22E-03 | 3,19E-03 | 2,94E-03 | 3,45E-03 | 1,17 | 1,08 | 1,26 |
| | 90 | 276 | 2,56E-03 | 2,54E-03 | 2,36E-03 | 2,75E-03 | 0,93 | 0,86 | 1,01 |

**Table S8: Days to percolation threshold by calendar start date (1 Jan–26 Dec).** Median and interquartile range (Q25–Q75) of days until the largest strong/weak component reaches n/2 of n connected wards, evaluated over N = 360 start dates. Delay is the maximum allowed gap (days) when forming temporal links; censored counts are number of start dates that did not reach the threshold by day 366.

| Strong components | | | | | | |
|---|---|---|---|---|---|---|
| Network | Delay (days) | N start dates | N Censored | median Cp (days) | Q25 | Q75 |
| All patients | 0 | 360 | 6 | 4 | 3 | 5 |
| | 14 | 360 | 0 | 1 | 1 | 2 |
| | 30 | 360 | 0 | 1 | 1 | 2 |
| | 90 | 360 | 0 | 1 | 1 | 2 |
| Inpatients | 0 | 360 | 9 | 7 | 6 | 8 |
| | 14 | 360 | 7 | 5 | 5 | 6 |
| | 30 | 360 | 5 | 5 | 4 | 5 |
| | 90 | 360 | 4 | 4 | 3 | 4 |
| **Weak components** | | | | | | |
| Network | Delay (days) | N start dates | Censored | median Cp (days) | Q25 | Q75 |
| All patients | 0 | 360 | 1 | 2 | 1 | 2 |
| | 14 | 360 | 0 | 1 | 1 | 2 |
| | 30 | 360 | 0 | 1 | 1 | 2 |
| | 90 | 360 | 0 | 1 | 1 | 1 |
| Inpatients | 0 | 360 | 2 | 2 | 2 | 3 |
| | 14 | 360 | 0 | 2 | 2 | 2 |
| | 30 | 360 | 0 | 2 | 1 | 2 |
| | 90 | 360 | 0 | 1 | 1 | 2 |

**Table S9: Distribution of proportions of patient movements in the aggregated hospital network, stratified by network type, flow direction, and movement component.** For each category, the table reports the number of hospitals contributing observations, N_hosp, the median proportion, first and third quartiles (q1, q3), interquartile range (IQR), and the 10th and 90th percentiles (p10, p90). Proportions represent the share of all patient movements originating from or terminating at each hospital that fall into the specified component.

| Network | Direction | Component | N_hosp | median | q1 | q3 | IQR | p10 | p90 |
|---|---|---|---|---|---|---|---|---|---|
| All patients | Outflow | Internal | 31 | 0,023 | 0,015 | 0,027 | 0,012 | 0,006 | 0,038 |
| | | Other hospitals | 36 | 0,013 | 0,009 | 0,02 | 0,011 | 0,007 | 0,041 |
| | | Outside | 36 | 0,961 | 0,954 | 0,973 | 0,019 | 0,937 | 0,985 |
| | Inflow | Internal | 31 | 0,023 | 0,015 | 0,027 | 0,012 | 0,006 | 0,038 |
| | | Other hospitals | 36 | 0,013 | 0,01 | 0,023 | 0,014 | 0,007 | 0,049 |
| | | Outside | 36 | 0,96 | 0,949 | 0,975 | 0,026 | 0,934 | 0,989 |
| Inpatients | Outflow | Internal | 30 | 0,036 | 0,011 | 0,05 | 0,039 | 0,002 | 0,069 |
| | | Other hospitals | 34 | 0,05 | 0,038 | 0,064 | 0,026 | 0,027 | 0,113 |
| | | Outside | 34 | 0,916 | 0,893 | 0,936 | 0,043 | 0,83 | 0,965 |
| | Inflow | Internal | 30 | 0,036 | 0,011 | 0,05 | 0,039 | 0,002 | 0,069 |
| | | Other hospitals | 34 | 0,054 | 0,042 | 0,089 | 0,048 | 0,028 | 0,118 |
| | | Outside | 34 | 0,906 | 0,882 | 0,92 | 0,038 | 0,819 | 0,968 |

#### Supplementary figures

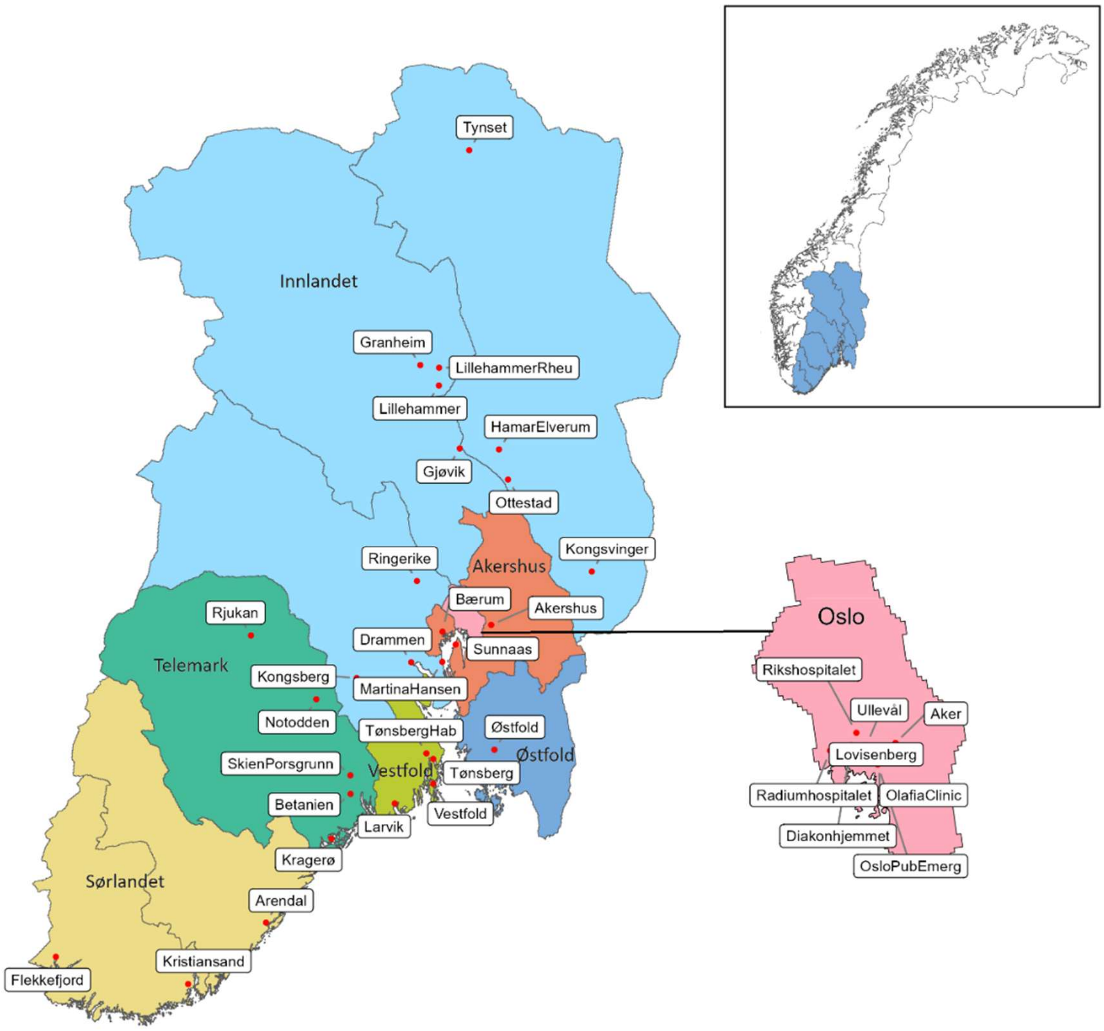


**S1 Fig. Map of South-Eastern Norway Regional Health Authority (HSØ) with location of the 36 hospitals and advanced treatment facilities in the region.**

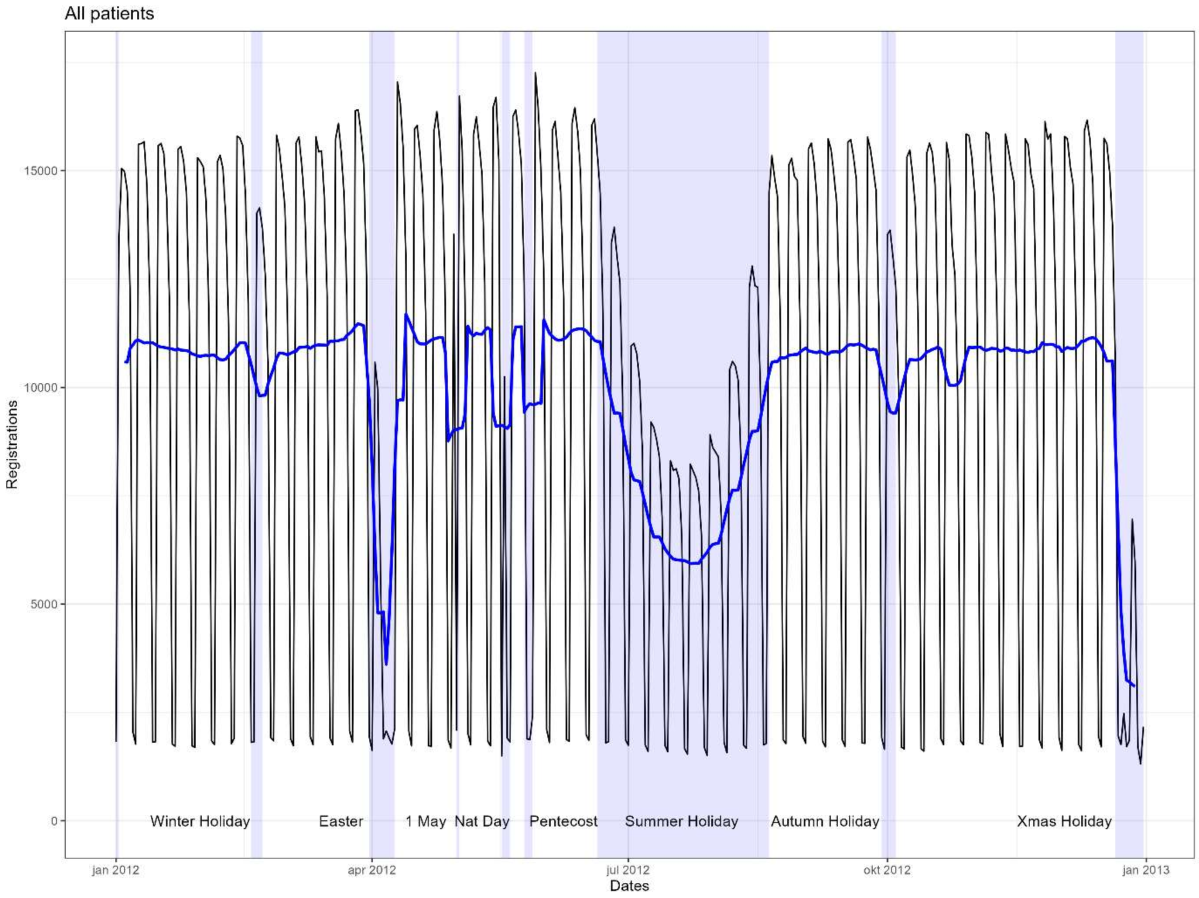


**S2A Fig. Daily counts of patient registrations in 2012 across all care types (inpatient, daycare, outpatient).** The black line shows the total number of registrations per calendar day, and the blue line a 7-day running average to smooth weekly fluctuations. Shaded blue vertical bars indicate Norwegian national and school holidays (Winter Holiday, Easter, Labour Day, Pentecost, Summer Holiday, Autumn Holiday, and Christmas). The plot highlights strong weekly cycles, with marked dips during summer and Christmas.

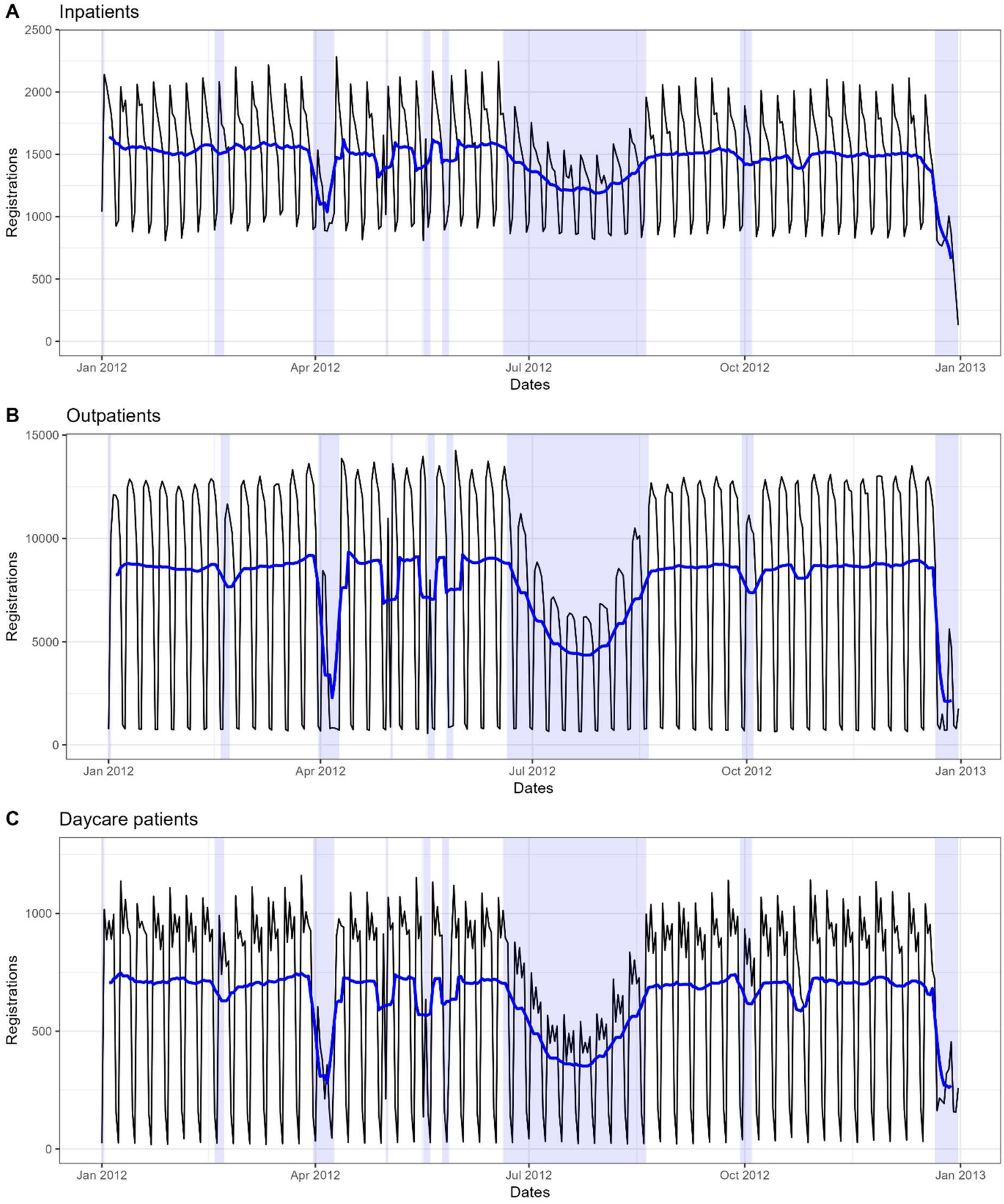


**S2B Fig Daily counts of patient registrations in 2012 by care type.** (A) Inpatients; (B) outpatients; (C) daycare patients. The black line shows total registrations per calendar day, and the blue line shows the 7-day running average to smooth weekly fluctuations. Shaded blue areas indicate national and school holidays. Outpatient visits account for ~80% of all registrations, with inpatients representing ~15%. Inpatient activity is relatively stable, decreasing by about half on weekends, whereas outpatient and daycare registrations drop sharply. Inpatient and daycare activity typically peak early in the week, while outpatient registrations peak mid-week.

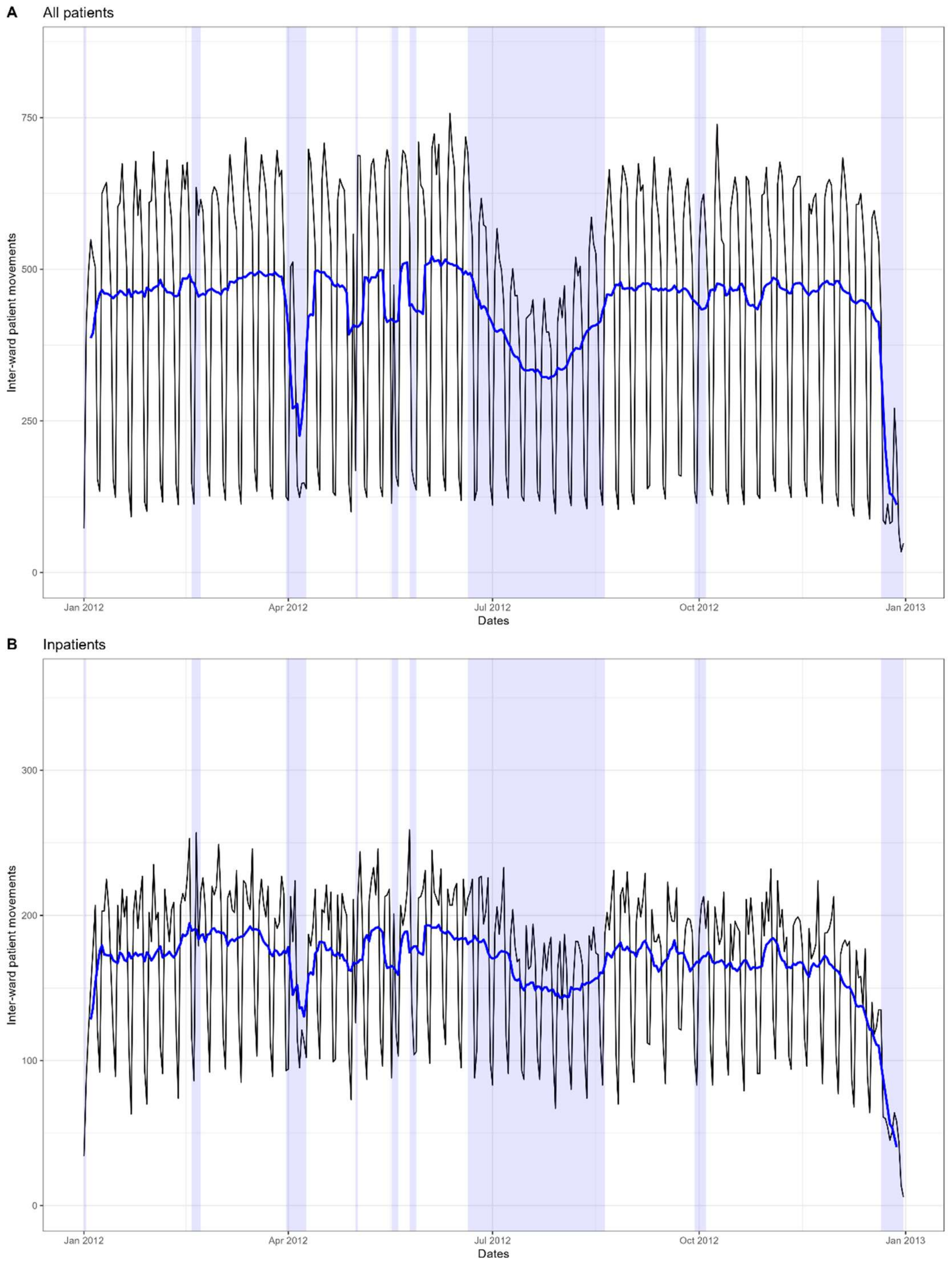


*a*

**Fig. Daily counts of inter-ward patient movements.** (A) All patients; (B) inpatients only. Patient movements exhibit clear weekly and seasonal patterns, with pronounced decreases on weekends and national holidays, mirroring trends in patient registrations. On weekdays, approximately 600 patients are transferred between wards, decreasing to around 100 on weekends, which are primarily inpatients. Inpatient transfer patterns are less variable, likely reflecting a higher degree of urgency and unplanned care. Censoring becomes apparent in early December for inpatients, reflecting longer treatment pathways, and later when outpatient and daycare patients are included.

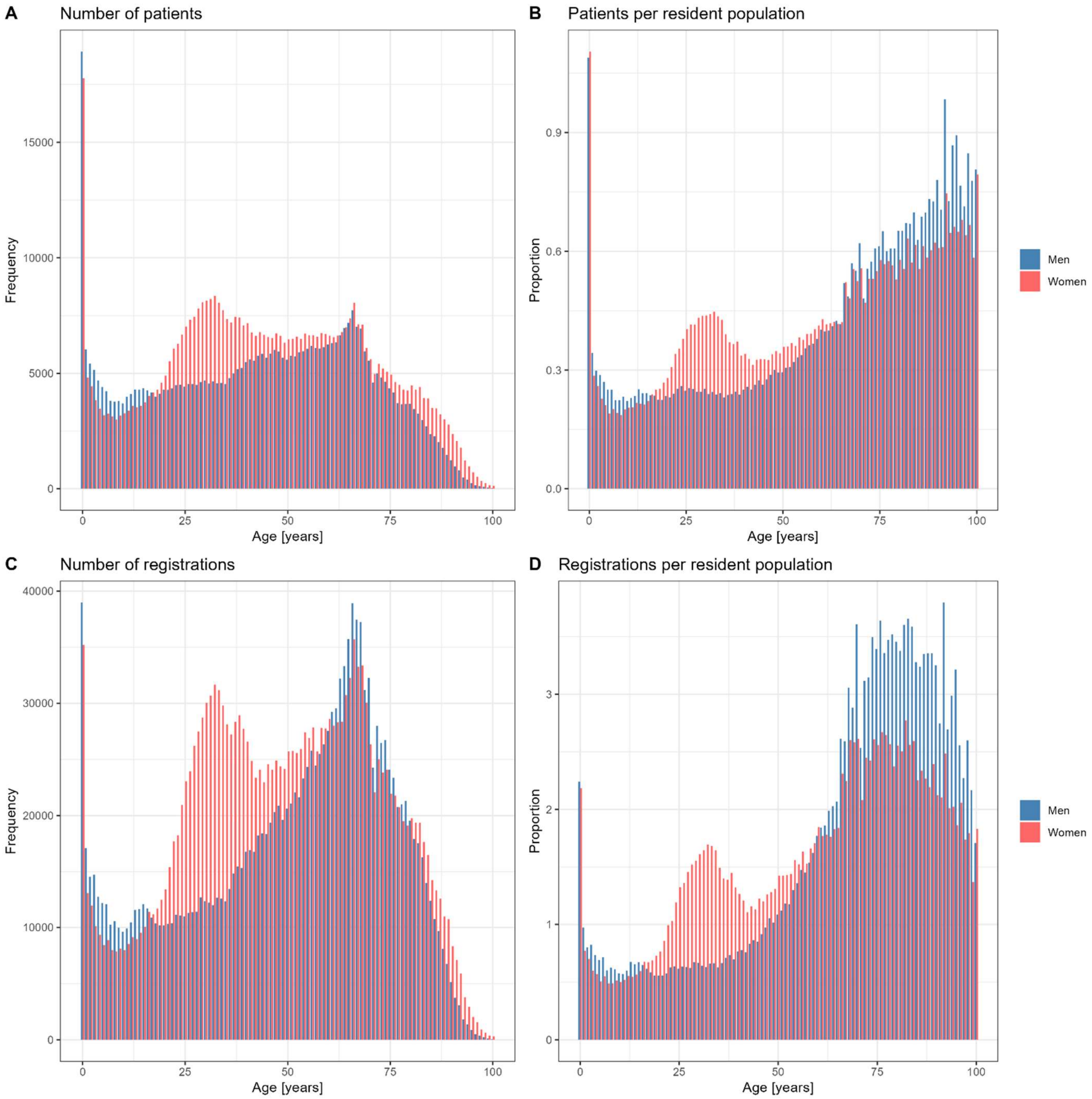


**Fig. S4. Age-specific distribution of hospital healthcare utilisation by sex.** (Men, blue; women, red). (A) Absolute number of patients; (B) number of patients per resident population. (C) Absolute number of registrations; (D) number of registrations per resident population.
Population-based measures are normalised by the age- and sex-specific resident population of the HSØ region in 2012. Note that population-normalised rates include both resident and non-resident patients in the numerator and therefore do not constitute a simple population risk measure.

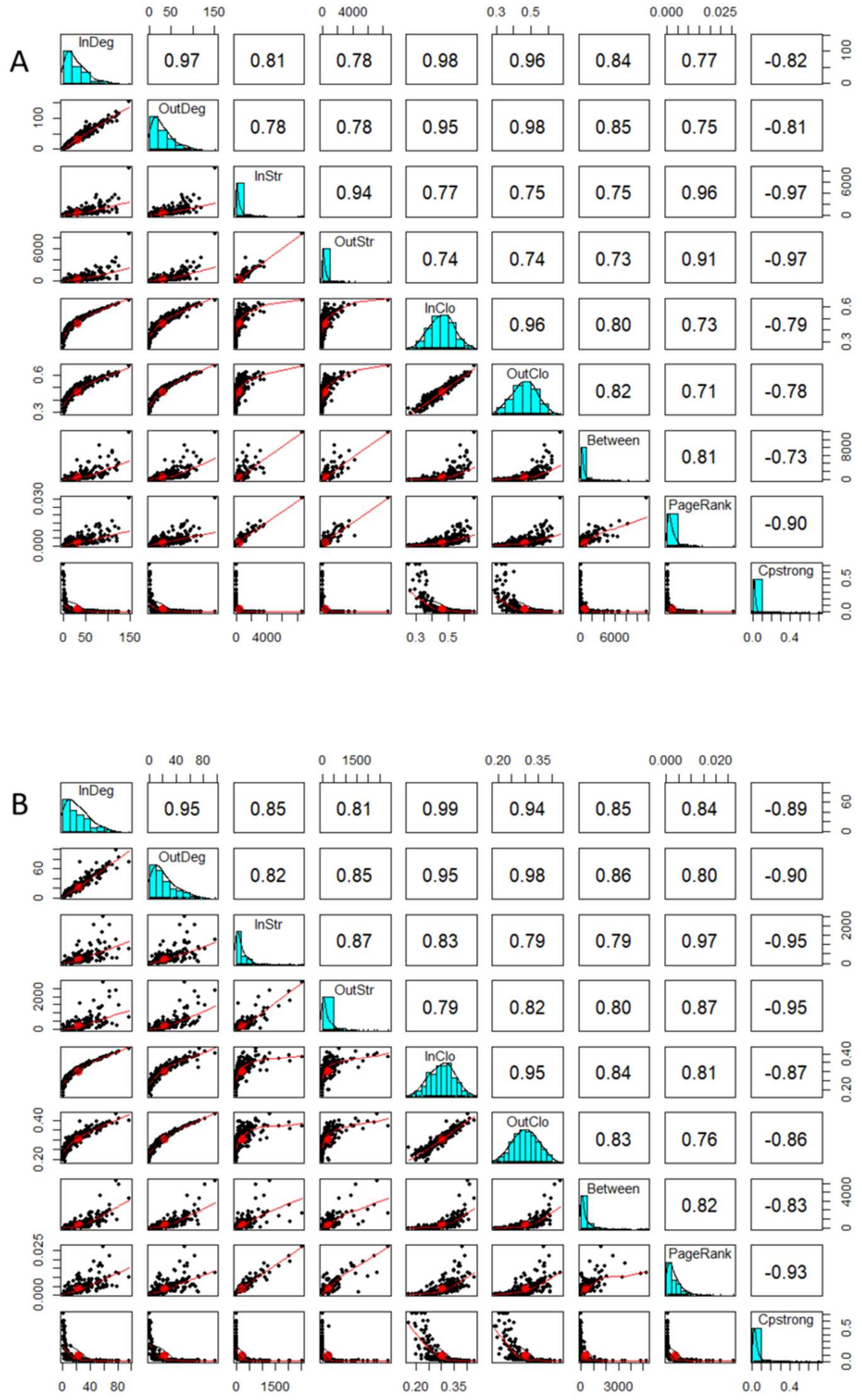


**S5 Fig. Pairwise correlations among ward-level centrality measures and percolation threshold $C_p$.** (A) All-patient network; (B) inpatient network. Each panel compares two metrics across wards. The upper triangle shows Spearman rank correlations ($\rho$); the diagonal shows marginal histograms; and the lower triangle shows scatterplots with least-squares trend lines (red). Measures include in- and out-degree, in- and out-strength, in- and out-closeness, betweenness, PageRank ($\alpha = 0.85$), and $C_p$(strongly connected components). Lower $C_p$values indicate higher structural robustness, leading to negative correlations with centrality measures.

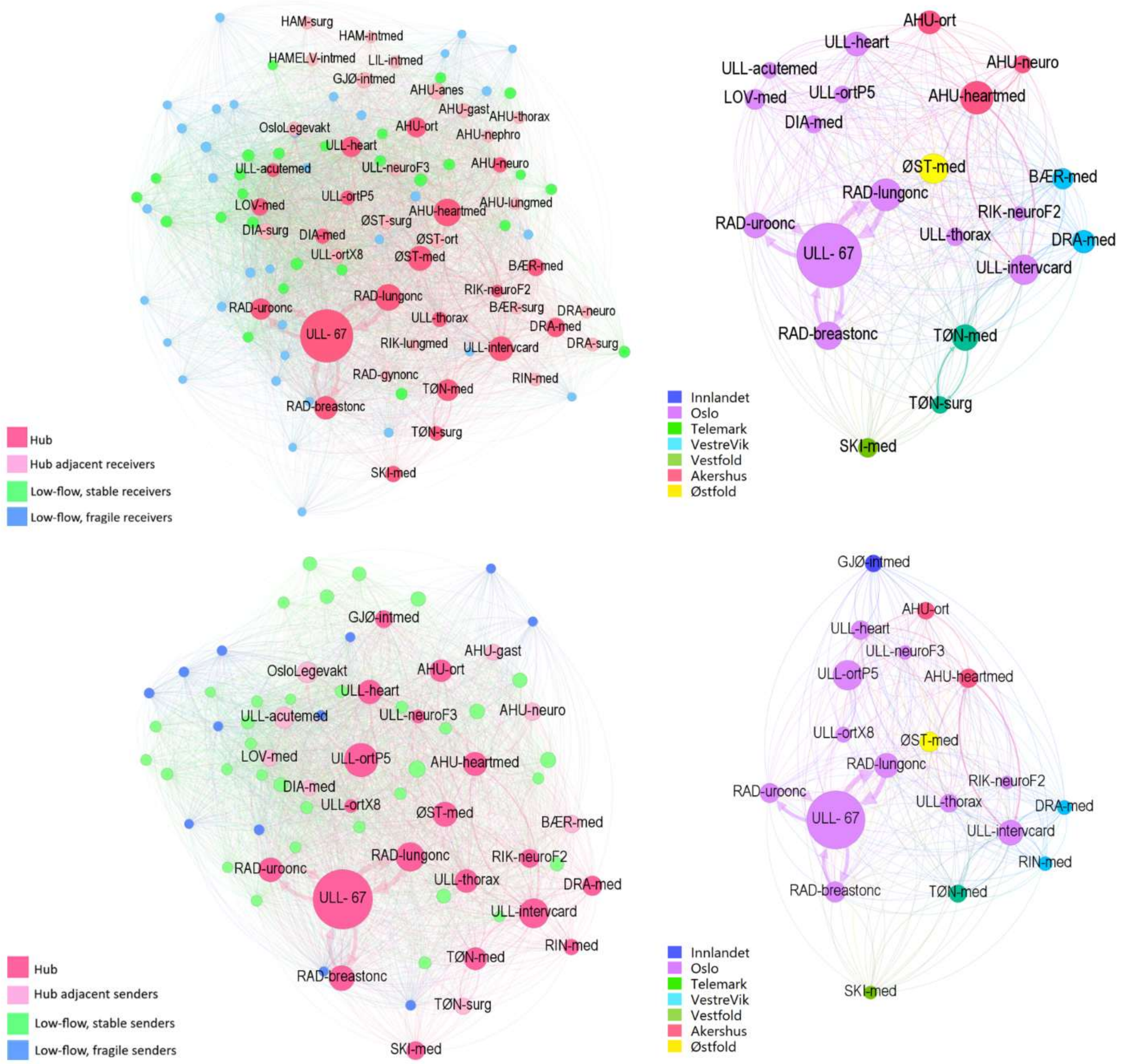


**S6 Fig. All-patient network central K-cores: roles and regional affiliation.** *(*A) Inflow central K-core showing GMM-derived ward roles; hub and hub–intermediary wards are labelled; (B) Hub and hub-intermediary wards in the inflow central K-core, coloured by region; (C) Outflow central K-core showing GMM-derived ward roles; hub and hub–intermediary wards are labelled; (D) Hub and hub-intermediary wards in the outflow K-core, coloured by region. Node size reflects in-strength (A–B) and out-strength (C–D). Note that wards may also maintain additional links outside the central K-core.

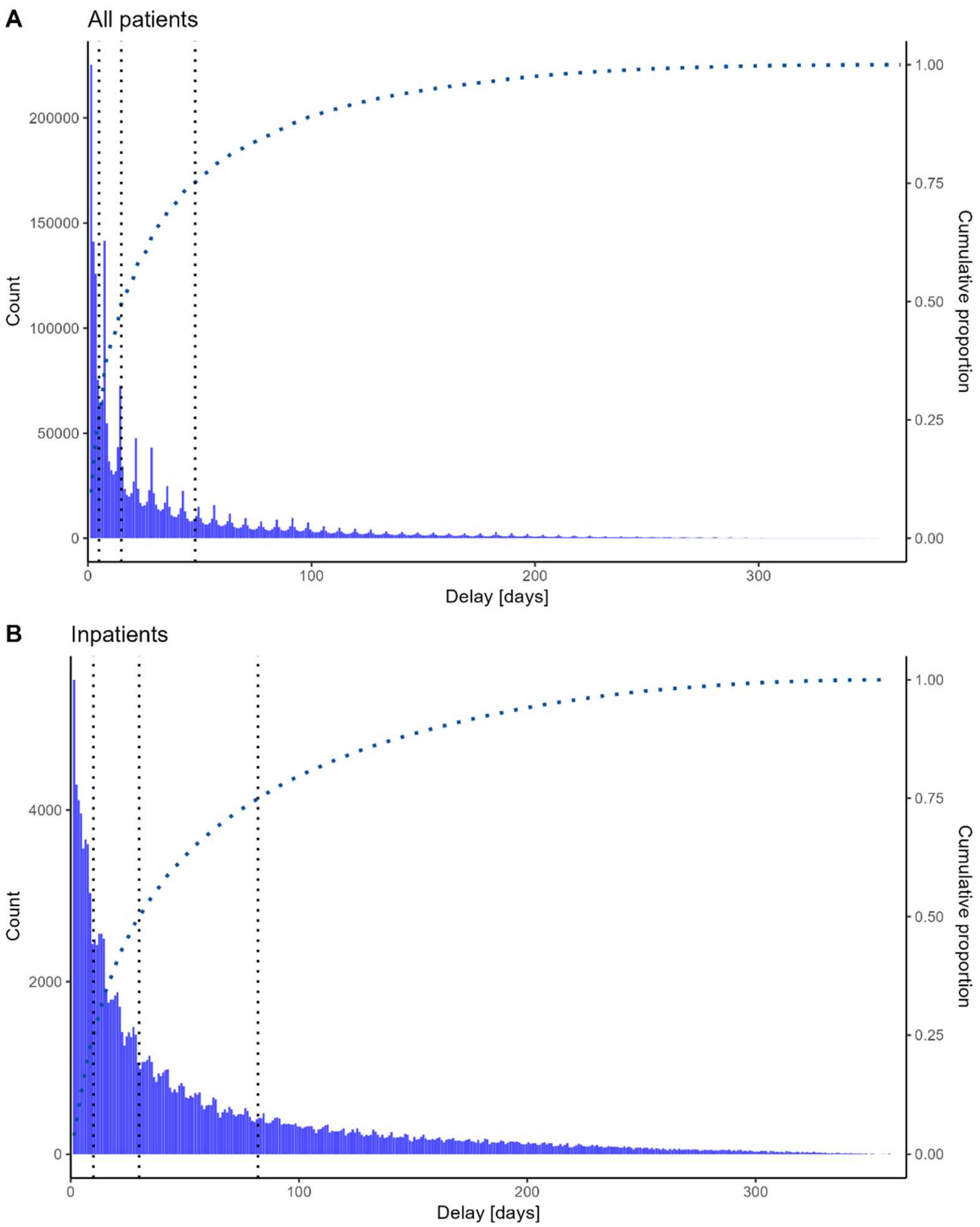


**S7 Fig. Empirical distribution of readmission delays and cumulative proportions.** (A) All-patient network ($\mathrm{N} = 2{,}338{,}390$); (B) inpatient network ($\mathrm{N} = 141{,}202$).

Blue bars show daily counts of inter-episode delays, and the dark blue dotted line the cumulative distribution (right axis). Vertical dotted lines indicate the 25th, 50th, and 75th percentiles. The all-patient network shows a strongly right-skewed distribution with pronounced weekly peaks, consistent with regular, planned attendances (e.g. dialysis, infusion, chemotherapy, and other recurring outpatient care). A secondary shoulder around ~30 days suggests monthly follow-up. In contrast, the inpatient distribution is broader, less sharply peaked at short delays, and lower in volume, accounting for 5.7% of all readmissions.

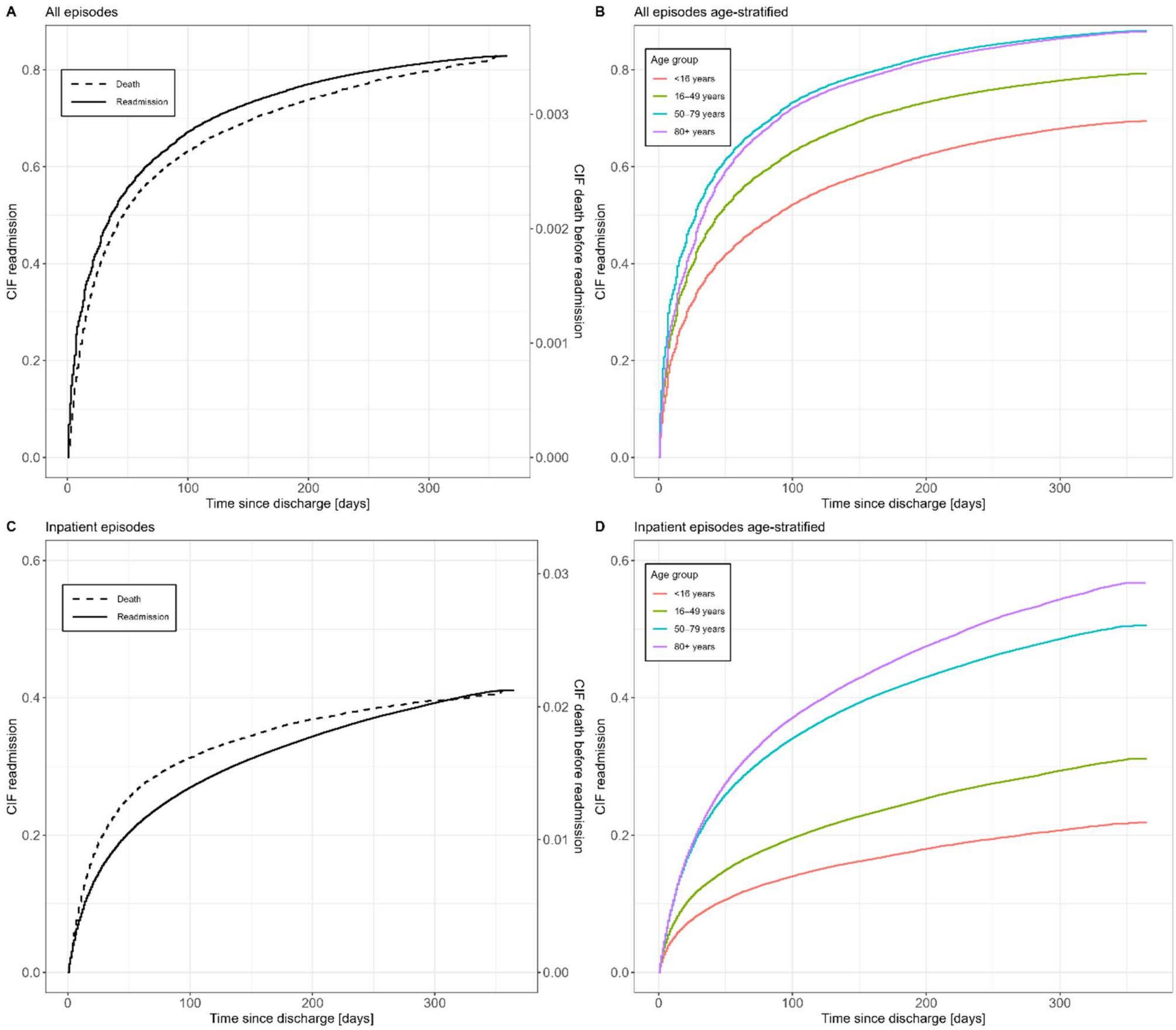


**S8 Fig. Cumulative incidence functions (CIFs) of readmission after hospital discharge, accounting for the competing risk of death before readmission, with episodes as the unit of analysis.** (A) All episodes (solid lines, readmission; stippled lines, death before readmission). (B) Age-stratified cumulative incidence curves for all episodes. (C) Inpatient episodes (solid lines, readmission; stippled lines, death before readmission). (D) Age-stratified cumulative incidence curves for inpatient episodes. Readmission accumulated rapidly after discharge. Death before readmission constituted a substantially more important competing event among inpatient than among all episodes. Readmission CIFs were generally higher in older age groups, with the clearest age gradient observed among inpatient episodes. Death before readmission was treated as a competing event, and episodes without readmission or death within 366 days were administratively censored. Episodes discharged on day 366 (3,284 all episodes; 1,326 inpatient episodes), episodes with admission recorded after the date of death (81 and 7 episodes, respectively), and episodes ending in death during the index hospitalisation (7,730 and 7,644 episodes, respectively) were excluded. The final analyses included 3,319,462 all episodes and 462,462 inpatient episodes. The flattening of the cumulative incidence curves towards the end of follow-up reflects increasing truncation of longer readmission intervals imposed by the one-year observation period (see S7 Fig).

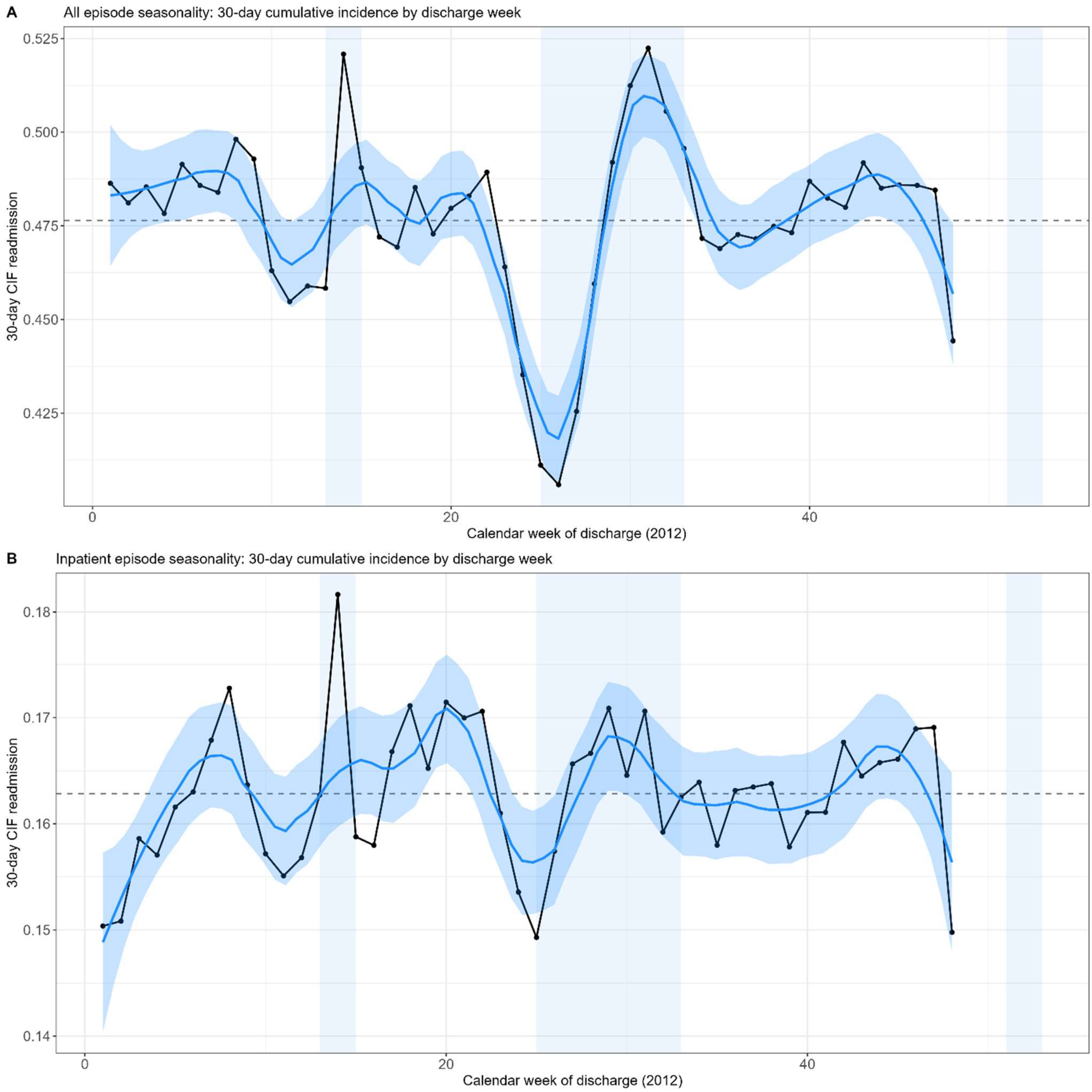


**S9 Fig. Weekly variation in the 30-day cumulative incidence function (CIF) of readmission after hospital discharge, accounting for the competing risk of death before readmission.** (A) All episodes. (B) Inpatient episodes. Black points connected by lines represent the estimated 30-day CIF of readmission for episodes discharged in each calendar week of 2012. The blue curve shows a LOESS-smoothed trend (span = 25%) with shaded 95% confidence bands. Horizontal dashed lines indicate the overall mean 30-day CIF across weeks. Vertical shaded areas denote holiday periods (Easter, summer holiday, and Christmas/New Year). Episodes discharged after calendar week 48 were excluded to ensure complete 30-day follow-up. The all-episode analysis suggests a transient reduction in the 30-day CIF of readmission in the period leading up to the summer holiday, followed by a rebound during the holidays; a similar but less pronounced temporal pattern is visible for inpatient episodes.
